# Selection of Mutations in HIV-1 Nucleocapsid and Integrase in Individuals Living with HIV Experiencing Virologic Failure After Initiating or Switching to Tenofovir-Lamivudine-Dolutegravir

**DOI:** 10.64898/2026.07.02.26356352

**Authors:** Kerri J. Penrose, Yuta Hikichi, Rahil Sethi, B. Jay Goetz, Lauren O. Siffert, Sujith A. Valiyaparmbil, Carole L. Wallis, Caitlyn McCarthy, Elizabeth Woolley, Uma R. Chandran, Cissy M. Kityo, Charles Flexner, Michael D. Hughes, Serena P. Koenig, Eric O. Freed, John W. Mellors, Urvi M. Parikh, the ACTG A5381-Hakim Study Team

## Abstract

**Background:** Tenofovir-lamivudine-dolutegravir (TLD) is an effective, single-tablet ART regimen but the causes of virologic failure that can occur despite adherence to TLD are incompletely understood, especially when resistance mutations in integrase (IN) are absent. Noncanonical resistance mutations in nucleocapsid (NC) have been selected in cell culture and are associated with decreased dolutegravir (DTG) susceptibility *in vitro*. Samples from the ACTG A5381-Hakim study were examined to assess the potential contribution of NC mutations to virologic failure on TLD.

**Methods:** A5381-Hakim was an observational cohort study that enrolled individuals with HIV-1 RNA >1000 copies/mL when initiating TLD as first line ART or switching from failing non-nucleoside reverse transcriptase inhibitor (NNRTI)-based or protease inhibitor (PI)-based ART. Whole HIV RNA genome next generation sequencing was performed on paired plasma samples from study entry and confirmed virologic failure in 56 participants receiving TLD for ≥six months. Mutations relative to the HIV_HXB2_ reference genome were identified with DeepChek® software using a mutation reporting threshold of five percent mixed-base frequency. Canonical drug resistance mutations (DRMs) were identified within the DeepChek® software using Stanford’s HIVdb v9.8 algorithm. Site-directed NC and IN mutants were tested for susceptibility to DTG, raltegravir and cabotegravir in the TZM-bl HIV-1 indicator cell line and evaluated for infectivity and replication in multi-round assays.

**Results:** Mutations in NC that emerged between TLD initiation and virologic failure were identified in 11 of 56 (20%) participants. Of these 11, three participants also had IN mutations at virologic failure but not at study entry, suggesting dual selection. Selected mutations in the NC zinc-finger domain included V13I, K20R, E21V, N27I/S, A30T, K34R, K38R, K41G/R/N, Q45R and/or M46I, alone or in combination with other NC and/or IN mutations. Specific NC mutations (K20R, N27I, A30T, K41N, M46I) conferred a significant decrease in DTG susceptibility. The combination of certain NC with IN mutations (e.g. NC N27I and IN R263K) conferred greater reduction in susceptibility to DTG *in vitro* than either mutation alone.

**Conclusions:** This work provides the first clinical evidence of NC mutation selection in individuals on failing TLD ART and shows that NC mutations, combined with IN mutations, can further decrease susceptibility to DTG. Our findings support additional investigations of the contributions of mutations outside of IN to virologic failure of integrase inhibitor-containing ART regimens.

## INTRODUCTION

The once-daily, fixed-dose tablet-containing tenofovir/lamivudine/dolutegravir (TLD) is a cornerstone of antiretroviral therapy (ART) in a majority of countries globally, with over 80% of people living with HIV-1 (PWH) taking dolutegravir (DTG)-based regimens due to high efficacy, lower cost (<45 U.S. dollars per person per year) and expanded distribution of generic products. ^1^ The World Health Organization recommends TLD for individuals initiating ART or switching from first-line non-nucleoside reverse transcriptase inhibitor (NNRTI)-based ART or second-line protease inhibitor (PI)-based ART. ^2^ Systemic reviews and meta-analyses that included over 6,000 participants have reported that ART-naïve individuals starting DTG-based ART are more likely to achieve viral suppression with fewer treatment discontinuations compared to individuals on NNRTI- and PI-based ART. ^3–5^ Both virologically suppressed and viremic individuals may benefit from switching to TLD as a second- or third-line regimen; however, higher pre-switch HIV-1 RNA levels, adherence challenges and pre-existing NNRTI resistance can increase the risk of DTG resistance in those who fail to maintain viral suppression. ^6–9^

Selection of HIV-1 integrase (IN) mutations that reduce susceptibility to integrase strand transfer inhibitors (INSTI) can contribute to virologic failure (VF) of DTG-based ART. Among PWH with VF on DTG-based ART, the proportion with INSTI drug resistance has ranged from 0% - 22.6%, with higher prevalence observed in ART-experienced individuals switching to TLD than in ART-naïve individuals initiating TLD. ^10–12^ The most common emergent IN mutations during VF in ART-experienced individuals include mutations at residues G118, E138, G140, and Q148, whereas R263K, G118R, N155H, or Q148H/R/K have been most commonly selected with VF in ART-naïve individuals. ^13,14^ Further, pre-existing IN mutations in INSTI-experienced individuals have been shown to lower response to DTG-containing ART in the VIKING 3 trial with increased likelihood of VF in individuals who have pre-existing Q148H/K/R with or without additional IN mutations. ^15^ Phenotypic susceptibility testing has shown variable resistance levels for the most common mutations selected during VF on INSTI-based regimens. Several INSTI resistance pathways have been shown to confer high-level phenotypic resistance to DTG, such as G118R alone or in combination with E138K. The R263K mutation, however, is more frequently detected, occurring in 40-50% of individuals who experience virologic failure on DTG-containing ART. ^12,14,16–18^ Although R263K is considered a major, non-polymorphic INSTI mutation, it causes only a modest two-fold reduction in susceptibility to DTG, bictegravir and cabotegravir, and HIV-1 variants with R263K have diminished replication fitness. ^19–22^

Studies of VF on TLD have shown that resistance mutations in IN may or may not be present. Inadequate ART adherence can explain VF in many but not all cases. ^23^ The mechanism of VF in those without IN resistance mutations is not well defined, but emerging evidence supports the potential role of non-canonical mutations outside of IN contributing to DTG resistance and VF. ^24^ For example, mutations in the 3’ polypurine tract (3’ PPT) have been selected by DTG *in vitro* and can lead to breakthrough viral replication without proviral integration in cell culture studies. ^25–28^ Additionally, nucleocapsid (NC) and envelope (Env) mutations selected under DTG pressure *in vitro* have been found to reduce susceptibility to DTG. NC mutations act by shortening the time between completion of reverse transcription and integration, thereby narrowing the window during which INSTIs can block integration. ^29^ Env mutations can reduce susceptibility to INSTIs by increasing viral replication capacity and enhancing cell-cell viral transmission. While Env mutations appear to have a broad impact on antiretroviral susceptibility *in vitro,* affecting all classes of ART, the contribution of NC mutations appears to be specific to INSTIs. ^24,29–31^ Detection of mutations in NC has recently been reported in individuals experiencing VF on DTG-containing ART in France, Cameroon, and in the UTRA study in South Africa, although samples before initiation of DTG-containing ART were not studied, leading to uncertainty about whether NC mutations were selected by therapy. ^32–35^

The Advancing Clinical Therapeutics Globally (ACTG) A5381-Hakim study was a prospective observational cohort designed to evaluate the therapeutic effectiveness of TLD and the emergence of HIV-1 drug resistance following initiation of TLD for first-, second- or third-line ART. ^23,36^ Primary findings were that participants who switched to TLD from failing first- or second-line ART achieved improved but suboptimal (<90%) viral suppression, with viral suppression rates that did not improve over time. Tenofovir diphosphate (TFV-DP) concentrations were lower among participants who remained unsuppressed compared with those who achieved virologic suppression, suggesting that incomplete adherence to TLD was a contributor to VF. ^23,36,37^ Across study groups, DTG-associated resistance mutations in IN were infrequently selected as assessed by Sanger sequencing. The study reported here sought to determine the potential contribution of non-canonical NC mutations to VF by analyzing carefully selected sample pairs from before switching to TLD and at VF on TLD among participants in the A5381-Hakim study.

## METHODS

### Study Design

A5381-Hakim enrolled PWH aged 10 years and older initiating TLD as first- or second-line ART through President’s Emergency Plan for AIDS Relief (PEPFAR)-supported HIV treatment programs at thirteen ACTG/PEPFAR-supported sites in six countries (South Africa, Malawi, Zimbabwe, Uganda, Kenya, and Haiti) between October 2019 and September 2022, with final sample collections in March 2023. The study was approved by local ethics committees and national regulatory agencies in the respective countries and registered with ClinicalTrials.gov, NCT04050449 **(Supplementary Table S1)**. ^23^ Written informed consent was obtained from all participants; minors provided assent along with parental or guardian consent.

### Study Samples

The analysis population included A5381-Hakim participants who were not virologically suppressed (HIV-1 RNA >1000 copies/mL) at study entry prior to the initiation of TLD and experienced confirmed VF, defined as two consecutive HIV-1 RNA measurements >1000 copies/mL (c/mL) occurring ≥six months after initiating TLD. Matched study entry (pre-TLD) and confirmed VF (post-TLD) plasma samples were obtained from A5381-Hakim Group 1a (individuals who switched from NNRTI-based first-line ART to TLD), Group 2a (individuals who switched from PI-based second-line ART to TLD), Group 3 (individuals who received first-line TLD with concomitant rifampicin treatment plus an additional 50 mg DTG), and Group 4 (individuals who initiated first-line TLD) **(Figure 1)**. Confirmed VF samples were collected at the second of the two consecutive follow-up visits with plasma HIV-1 RNA >1000 c/mL, which could have occurred up to six months after the first visit with HIV-1 RNA >1000 c/mL during the next scheduled study visit.

**Figure 1.**
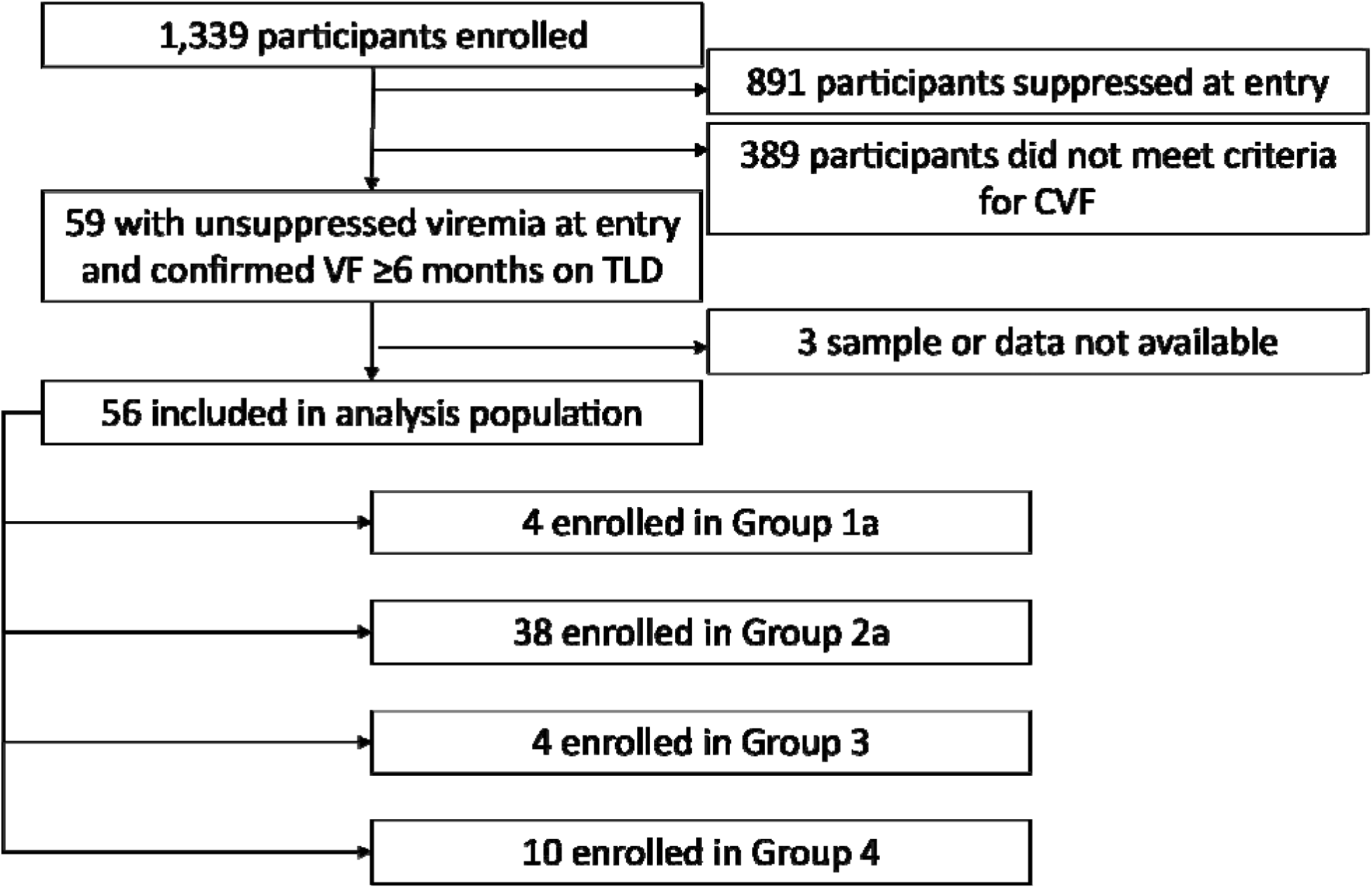
STROBE Diagram. Analysis population includes participants with viremia at study entry and confirmed virologic failure (CVF) defined as two consecutive HIV-1 RNA measures >1000 copies/mL (c/mL) on or after six months of initiating a single-tablet antiretroviral therapy (ART) regimen containing tenofovir/lamivudine/dolutegravir (TLD). In Group 1a, individuals switched to TLD from a non-nucleoside reverse transcriptase inhibitor (NNRTI)-containing regimen with an enrollment HIV-1 RNA >1000 c/mL. In Group 2a, individuals switched to TLD from a protease inhibitor (PI)-containing regimen with an enrollment HIV-1 RNA >1000 c/mL. In Group 3, individuals received first-line TLD with rifampin-containing tuberculosis treatment and an additional 50 mg dose of dolutegravir. In Group 4, treatment-naïve individuals initiated first-line TLD. Full details of the Hakim study population have been previously published. ^23,36^

### Study Procedures

HIV-1 RNA was monitored locally at study entry and at six, 12 and 24 months post-TLD initiation using standard-of-care assays. ^23,36^ Tenofovir diphosphate (TFV-DP) concentrations were measured at the University of Cape Town from dried blood spots collected at confirmed VF to approximate ART adherence, as previously described. ^38^ Briefly, a 25 µl DBS punch was extracted with methanol: water and isotopic tenofovir as an internal standard, then TFV-DP was quantified by validated liquid chromatography/tandem mass spectrometry. The lower limit of quantification was 16.6 femtomole (fmol)/3mm punch. ^39–41^ HIV-1 genotype analysis was performed on plasma specimens with HIV-1 RNA >1000 copies/mL at six months and confirmed VF by Sanger sequencing at BARC-South Africa/Lancet Laboratories or the Pittsburgh Virology Specialty Laboratory (University of Pittsburgh, Pennsylvania, USA). HIV-1 drug resistance mutations were identified using the Stanford HIVdb algorithm v8.9 as previously described. ^42–44^

### Whole HIV-1 RNA Genome Sequencing

A laboratory-developed whole HIV-1 RNA genome sequencing (HIV-1 WGS) method was used to identify mutations in NC, protease (PR), reverse transcriptase (RT) and IN at a variant frequency of ≥5%. Total RNA was extracted from plasma using guanidinium thiocyanate lysis followed by isopropanol precipitation. HIV-1 whole-genome Illumina libraries were prepared using an approach adapted from Gall *et al.* to amplify HIV-1 RNA genomes in four overlapping one-step RT-PCR reactions (Superscript IV One Step, ThermoFisher Scientific). ^45^ Primers were optimized for subtype diversity, optimal amplification across a range of HIV-1 RNA >1000 copies/mL and long terminal repeat (LTR) coverage through partial R and full U5 in amplicon one and full U3 and partial R in amplicon four. Optional nested PCR primers were used in a second round of PCR (SuperFI, ThermoFisher Scientific) for samples with poor amplification **(Supplementary Table S2)**. ^46^ Amplicons were purified using bead- or agarose gel-based purification (KAPA Pure Bead, Roche) and quantified by PicoGreen Quant-it (Thermo Fisher Scientific) prior to library preparation (PurePlex, SeqWell). Libraries were pooled, quantified, and sequenced on the Illumina MiSeq platform. Paired-end reads were demultiplexed using Illumina software, and FASTQ files were generated for downstream bioinformatic analysis.

### Bioinformatics

DeepChek® software using a mutation threshold of five percent frequency was used for analysis of all HIV-1 gene targets. Drug resistance mutations (DRMs) were identified by the DeepChek® software using Stanford’s HIVdb v9.8 algorithm ^47^. The frequency of each variant present at ≥5% was obtained from the Amino Acid - Quality Information Report. Mutations were considered selected at confirmed VF on TLD if they were detected at ≥20% frequency at confirmed VF and were either absent at study entry or present at <20% frequency at study entry.

### Phenotypic analysis of NC and IN Mutations

#### Plasmids

The full-length HIV-1 molecular clone pNL4-3 and nanoLuc reporter, pNL4-3 6ATRi-nanoLuc KFS were used for phenotype analyses of mutants and wild-type controls ^29,48^. Mutations found to be selected at the time of confirmed VF in the *gag* and *integrase* genes were introduced into pNL4-3–based plasmids by overlapping polymerase chain reaction (PCR) and site-directed mutagenesis. The VSV-G–expressing plasmid, pHCMVG, was a gift from J. Burns. ^49^

#### Cells and Reagents

Human embryonic kidney (HEK) 293T, and TZM-bl cells were maintained in Dulbecco’s modified Eagle’s medium supplemented with 10% fetal bovine serum (FBS) at 37°C in 5% CO_2_. ^50^ The SupT1 T-cell line was cultured in RPMI-1640 medium supplemented with 10% FBS at 37°C in 5% CO_2_. DTG and CAB were purchased from MedChemExpress and Cayman Chemical, respectively. Raltegravir (RAL) was obtained through the BEI Resources’ HIV Reagent Program. Human peripheral blood mononuclear cells (PBMCs) were obtained from anonymous, de-identified blood donors enrolled in the United States National Institutes of Health Department of Transfusion Medicine Blood Products Program (NIH CC-DTM). Purified primary CD4+ T-cells were isolated from PBMCs using EasySep Human CD4+ T-cell isolation kit (STEMCELL Technologies), followed by expansion of the cells using 50 Units/mL IL-2 and ImmunoCult™ Human CD3/CD28 T Cell Activator (STEMCELL Technologies) according to the manufacturer’s instructions. Three days before infection, the purified CD4+ T-cells were restimulated with 2 μg/mL phytohemagglutinin (PHA-P).

#### Preparation of virus stocks

HEK293T cells were transfected with HIV-1 plasmid DNA using Lipofectamine 2000 (Invitrogen) according to the manufacturer’s instructions. VSV-G–pseudotyped viruses were prepared by co-transfecting HEK293T cells with pNL4-3 6ATRi-nanoLuc KFS and pHCMVG at a DNA ratio of 10:1. At 48 hours post-transfection (26 hours for VSV-G-pseudotyped virus), virus-containing supernatants were filtered through a 0.45-μm membrane filter (Merck Millipore). The amount of virus in the supernatant was quantified by RT assay as previously described. ^24^

#### Single-round infectivity assay

Single-round infectivity assays were performed as previously described. ^24^ TZM-bl cells (1.0 x 10^4^ cells) in 96-well plates were incubated with RT-normalized virus inoculum in the presence of various concentrations of antiretroviral drugs with diethylaminoethyl (DEAE)-dextran (30 μg/mL). Infectivity assays were performed within the linear range of the assay. At the indicated time after infection, luciferase activity was measured using the Britelite plus reporter gene assay system (PerkinElmer) and GloMax Navigator microplate luminometer (Promega). PHA-stimulated primary CD4 T-cells (2.0 x 10^6^ cells) were incubated with RT-normalized VSV-G pseudotyped nanoLuc reporter virus at 37°C for 2 hours. Following incubation, the cells were washed and plated in 96-well flat-bottom plates and incubated at 37°C in the presence of various concentrations of DTG. Nano-luciferase activity was measured using Nano-Glo luciferase system (Promega) according to the manufacturer’s instructions.

#### Virus replication kinetics assays

Virus replication was monitored in SupT1 cells as previously described. ^24^ Briefly, SupT1 cells were incubated with the indicated pNL4-3 clones (1.0 μg of DNA/1.0 x 10^6^ cells) in the presence of DEAE-dextran (700 μg/mL) at 37°C for 15 min. Transfected cells (1.5 x 10^5^ cells) were plated in 96-well flat-bottom plates and incubated at 37°C in the presence of varying concentrations of DTG. Aliquots of supernatants were collected to measure RT activity, and cells were split 1:3 every other day with fresh drug and medium. IC_50_ values were calculated based on the area under the curve (AUC) of the replication kinetics. IC_50_ was defined as the amount of inhibitor required to reduce the AUC by 50%.

## RESULTS

### Participant Characteristics

Of 1,339 participants enrolled in A5381-Hakim, 891 who had HIV-1 RNA <1000 c/mL at study entry and 389 who had HIV-1 RNA >1000 copies/mL at study entry but did not meet the definition of confirmed VF after ≥6 months of TLD treatment were excluded from this analysis. Three participants did not have data and/or samples available. Therefore, the analysis population included 56 participants who started A5381-Hakim with unsuppressed viremia before initiating or switching to TLD and subsequently experienced confirmed VF after at least six months of TLD treatment **(Figure 1)**.

Participants were 54% female and a median of 31 years of age (min, max: 11, 68). Twenty participants (36%) were from Haiti, 14 (25%) from Kenya, 11 (20%) from Uganda, ten (18%) from Malawi, and one (2%) from South Africa (**Table 1)**. Participants had a median plasma HIV-1 RNA level of 40,235 copies/mL (IQR: 9,221-193,776) at study entry, and experienced confirmed VF a median of eleven months (IQR: 10 – 17) after initiating TLD. At the time of sampling for HIV-1 WGS, which occurred between one and seven months after the initial HIV-1 RNA measurement >1000 copies/mL, participants had a median HIV-1 RNA at confirmed VF of 29,973 copies/mL (IQR: 6,537 – 106,365).

**Table 1.** Participant Characteristics.

|  | **Analysis Population**<br>**(N = 56)** |
| --- | --- |
| Female Sex | 30 (54%) |
| **Age (years)** |  |
| Median (IQR)<sup>a</sup> | 31 (22.5, 43.5) |
| Min, Max | 11, 68 |
| **Country** |  |
| Haiti | 20 (36%) |
| Kenya | 14 (25%) |
| Uganda | 11 (20%) |
| Malawi | 10 (18%) |
| South Africa | 1 (2%) |
| **Viremia** |  |
| Median (IQR) HIV-1 RNA at Study Entry (c/mL)<sup>a</sup> | 40,235 (9,221-193,776) |
| Median (IQR) Time to Confirmed Virologic Failure, CVF (mo.)<sup>a,b</sup> | 11 (10-17) |
| Median (IQR) HIV-1 RNA at CVF (c/mL)<sup>a,b</sup> | 29,973 (6,537-106,365) |
| **TFV-DP Levels in DBS at CVF<sup>b,c</sup>** |  |
| No. with measurement | 51 (91%) |
| No. with TFV-DP levels ≥ LOQ | 31 of 51 (61%) |
| No. with TFV-DP levels < LOQ of 16.6 fmol/3 mm punch | 20 of 51 (39%) |
| Median (IQR) TFV-DP levels among those ≥ LOQ (fmol/3 mm punch) | 287 (89-397) |
| **Resistance Mutations in RT and/or IN** |  |
| No. with no mutations in RT and IN at Study Entry and CVF | 11 (20%) |
| No. with no mutations in RT and with mutations in IN at Study Entry and CVF | 1 (2%) |
| No. with mutations in RT and no mutations in IN at Study Entry | 36 (64%) |
| AND with mutations in RT and no mutations in IN at CVF | 25 of 36 (69%) |
| AND with mutations in RT and with mutations in IN at CVF | 5 of 36 (14%) |
| AND with no mutations in RT and no mutations in IN at CVF | 6 of 36 (17%) |
| No. with mutations in RT and IN at Study Entry | 8 (14%) |
| AND with mutations in RT and mutations in IN at CVF | 7 of 8 (88%) |
| AND with mutations in RT and no mutations in IN at CVF | 1 of 8 (13%) |

<sup>a</sup>. Interquartile range is reported as 25<sup>th</sup> percentile to 75<sup>th</sup> percentile.
<sup>b</sup>. Months from start of TLD to confirmed virological failure (CVF) is defined as two consecutive HIV-1 RNA measurements >1000 c/mL after at least six months of TLD treatment.
<sup>c</sup>. The limit of quantitation (LOQ) for detection of TFV-DP in DBS is 16.6 fmol/3mm punch.
Abbreviations: Number (No.); Interquartile range (IQR); protease (PR); reverse transcriptase (RT); integrase (IN); copies/milliliter plasma (c/mL); months (mo); number (no.); tenofovir diphosphate (TFV-DP); dried blood spots (DBS); limit of quantitation (LOQ); millimeter (mm); pharmacokinetics (PK); femtomole (fmol)

### Adherence Estimates by Levels of Detectable Tenofovir Diphosphate

TFV-DP data were available for 51 of 56 participants at the time of confirmed VF. Among these 51 participants, 31 (61%) had measurable TFV-DP above the lower limit of quantitation (LOQ) of 16.6 femtomole per 3 millimeter dried blood spot punch (fmol/3mm punch), with a median TFV-DP concentration of 287 (IQR: 89-397) fmol/3mm punch (**Table 1**).

### Resistance-Associated RT Mutations and Accessory IN Mutations in Individuals on Failing Tenofovir-Lamivudine-Dolutegravir Antiretroviral Therapy

Of the 56 participants, 11 (20%) had no mutations in RT and IN at both study entry and confirmed VF. Only 6 of these 11 had detectable levels of TFV-DP at confirmed VF. One participant (2%) had only IN-L74I present at both study entry and confirmed VF with no mutations in RT. Thirty-six of 56 (64%) participants had resistance mutations in RT at study entry; of these 36, 25 (69%) had no IN mutations at entry or confirmed VF and 5 (14%) had selected mutations in IN at confirmed VF that were not present at study entry, which included the major IN mutations G118R and R263K, and the accessory mutations IN-H51Y and IN-L74M. Eight participants had mutations in both RT and IN at study entry; seven of those eight had the same IN accessory mutations at both timepoints including L74I and/or E157Q. T97A was present in one participant at low frequency (9%) only at study entry **(Table 1).** Full genotypes and drug levels for each participant are presented in **Supplementary Table S3**.

### Selection of Resistance-Associated IN Mutations and NC Mutations in Individuals on Failing Tenofovir-Lamivudine-Dolutegravir Antiretroviral Therapy

As shown in **Figure 2**, only five of 56 individuals with confirmed VF on DTG-based ART had major and/or accessory resistance-associated mutations in IN selected at confirmed VF that were not present at study entry. Two had IN mutations only; these included 100% G118R with the H51Y accessory mutation at 5% frequency in PID16, and 100% G118R with the accessory mutation L74M at 28% frequency in PID33. Three participants had concurrent emergence of IN and NC mutations at confirmed VF, including PID 6 (41% NC-K41G; 62% IN-G118R), PID 21 (77% NC-N27I; 100% IN-R263K), and PID 27 (59% NC-A30T, 69% NC-M46I; 12% IN-H51Y). All five participants had complex PR/RT resistance profiles that were present at study entry and maintained at confirmed VF. TFV-DP levels were available for four of five individuals at confirmed VF, indicating moderate to high level of adherence to TLD (range of 329 – 1251 fmol/3mm punch).

**Figure 2.**
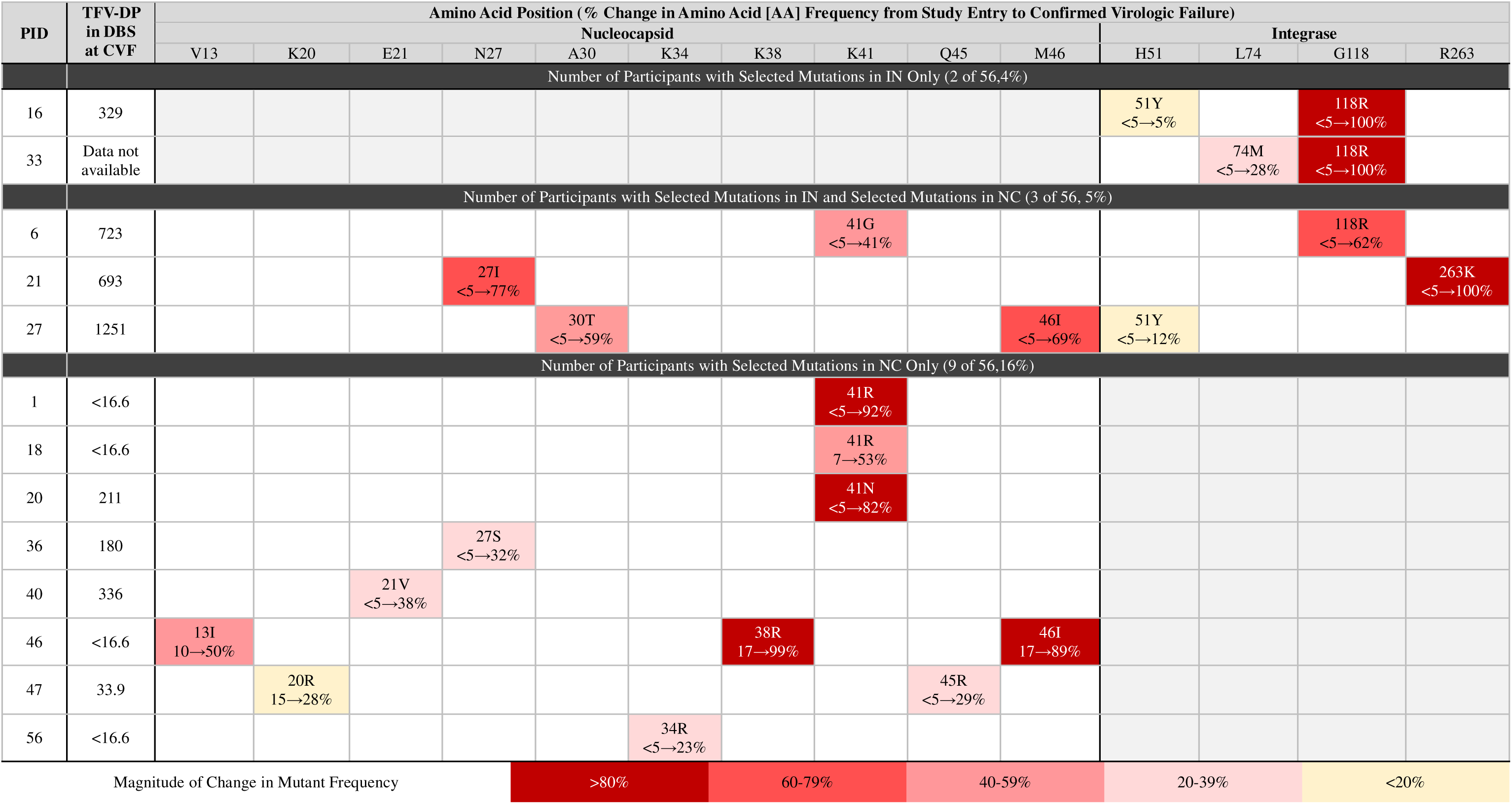
Major Resistance-Associated Integrase (IN) Mutations and Selected Nucleocapsid (NC) Mutations in Individuals on Failing Tenofovir-Lamivudine-Dolutegravir Antiretroviral Therapy. Limit of quantitation (LOQ) for detection of TFV-DP in DBS is 16.6 femtomole per three millimeter dried blood spot punch. Confirmed Virologic Failure (as defined in Methods) samples were collected at the second of the two consecutive follow-up visits with plasma HIV-1 RNA >1000 copies/mL. Shaded boxes indicate magnitude of change in mutant frequency. Abbreviations: Participant Identifier (PID); tenofovir diphosphate (TFV-DP); dried blood spot (DBS); confirmed virologic failure (CVF)

Eight individuals had selected mutations in the zinc finger motif region of NC without mutations in IN at confirmed VF. Two of the eight participants selected NC mutations at confirmed VF in the absence of RT and IN resistance mutations at entry, including NC-K41N alone (PID 20) and the combination of NC-V13I, NC-K38R and NC-M46I (PID 46). Six of the eight participants selected mutations in NC in a background of multiple RT mutations at entry (PIDs 1, 18, 36, 40, 47, and 56). Of these eight participants, four had undetectable TFV-DP at confirmed VF, and four had TFV-DP levels ranging from 180 – 1251 fmol/punch **(Figure 2 and Supplementary Table S3)**.

### Evaluation of 3’ Polypurine Tract (PPT)

Mutations in the 3’ polypurine tract have been previously implicated in INSTI drug resistance. ^51^ No mutations were found to be selected in the 3’ PPT at the time of confirmed VF.

### Phenotypic effects of NC mutations on INSTI resistance

To better understand the role of zinc-finger NC mutations in INSTI resistance, NC and/or IN mutations that emerged in study participants were introduced into HIV-1_NL4-3_ by site-directed mutagenesis and evaluated for susceptibility to DTG **(Figure 3A)**. Phenotypic evaluation of 10 unique NC mutations selected in individuals with confirmed VF demonstrated that five individual NC mutations - K20R, N27I, A30T, K41N and M46I - significantly decreased susceptibility to DTG (1.5 to 2.3-fold; p < 0.05) compared to wild-type (WT) HIV-1 **(Figure 3B)**. To evaluate if NC mutations also caused reductions in susceptibility to other INSTIs, these mutations were further evaluated against raltegravir (RAL) and cabotegravir (CAB). These same five mutations (K20R, N27I, A30T, K41N and M46I) as well as K41R also reduced susceptibility to RAL (1.5 to 1.8-fold; p < 0.05; **Figure 3D**). Reduction in susceptibility to CAB was conferred by N27I, A30T, K41N, Q45R or M46I (1.6 to 2.1-fold, p < 0.05; **Figure 3E**). The A30T/M46I double mutant (corresponding to the genotype detected in PID 27) exhibited the highest fold-decrease in susceptibility to all 3 inhibitors: DTG (2.9-fold; p < 0.05), RAL (2.4-fold; p < 0.05) and CAB (3.1-fold; p < 0.05) **(Table 2**, **Figures 3B, D, E).** Linear regression analysis of DTG IC_50_s for genotypes detected in study participants and an additional 15 genotypes selected *in vitro* by DTG from Hikichi *et al.* (2024) demonstrated significant correlations between IC_50_s of DTG and CAB and between DTG and RAL (**Figures 3F and G**).

**Figure 3.**
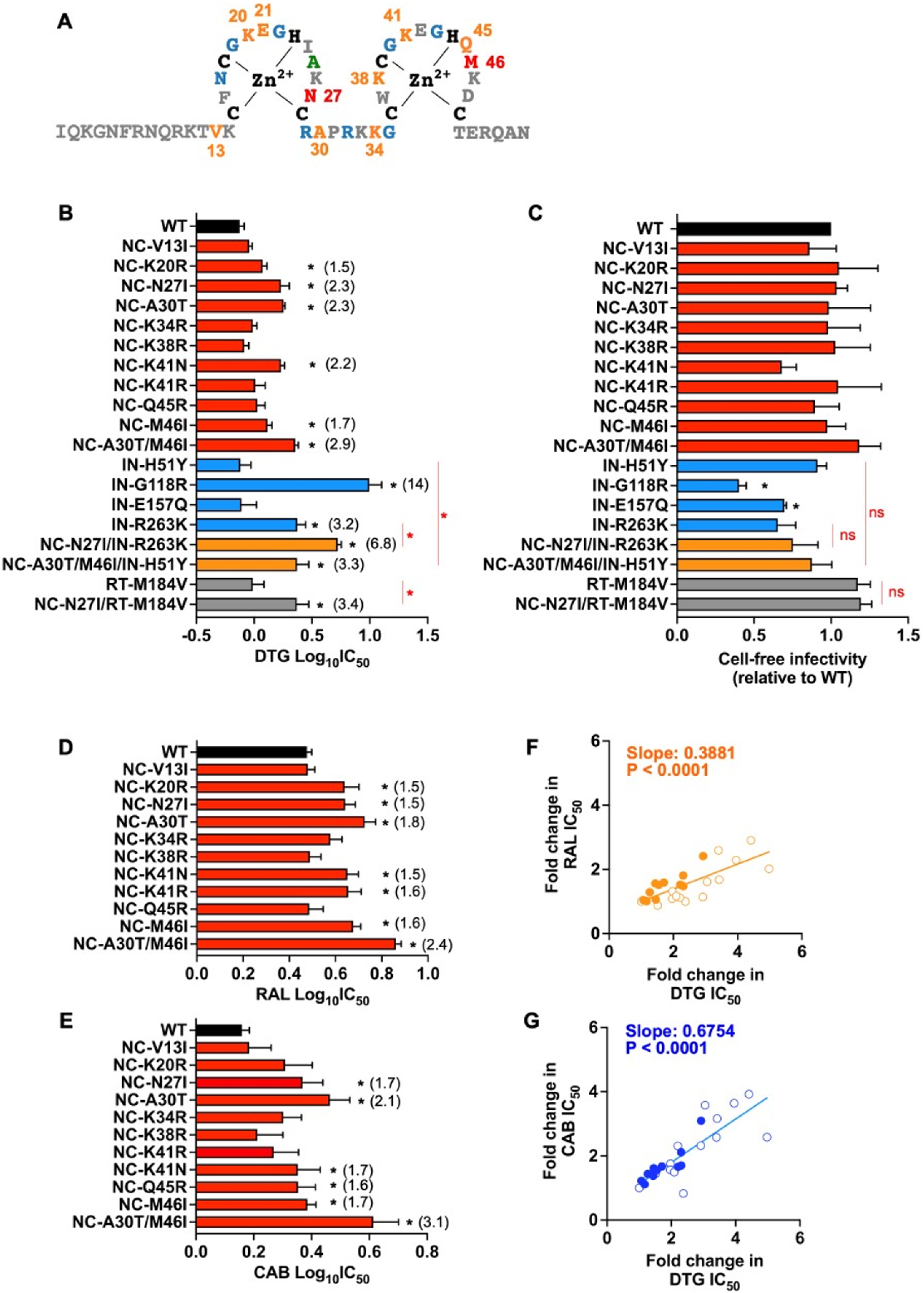
INSTI Susceptibility of HIV-1 with Clinically Observed Nucleocapsid and Integrase Mutants. (A) Sequence of the HIV-1 nucleocapsid (NC) domain. Integrase stand transfer inhibitor (INSTI)-resistance mutations identified in both *in vitro* dolutegravir (DTG) selection experiments as previously reported in Hikichi et al. and from A5381-Hakim study participants in the current analysis population are shown in red. ^24^ Mutations identified only *in vitro* selection experiments are shown in blue. Newly identified NC mutations from the current analysis population are shown in orange. Variants detected previously *in vitro* but not conferring drug resistance are shown in green. B) Susceptibility of mutants to DTG. TZM-bl cells were infected with reverse transcriptase (RT)-normalized wild-type HIV-1_NL4-3_ (WT), NC mutant (red), integrase (IN) mutant (orange), or NC + IN mutants (blue) in the presence of a range of concentrations of DTG (from 0.03 to 1000 nM DTG). Luciferase activity was measured 48lJhours after infection. RT-M184V was included as a nucleoside reverse transcriptase inhibitor (NRTI)-resistant control mutant (gray). Data from at least three independent experiments are shown as means ± standard error of the mean (SEM). \**P* < 0.05, one-way analysis of variance (ANOVA), followed by two-stage step-up method of Benjamini, Krieger, and Yekutieli (mutants vs WT) or Welch’s t-test (comparison of IN vs IN+NC mutants). ^56^ (C) Cell-free viral infectivity of NC mutants. RT-normalized virus stocks produced from 293T cells were used to infect TZM-bl cells. Luciferase activity was measured 48 hours after infection. The infectivity of HIV-1_NL4-3_ (WT) is normalized to 1.0. Data from at least three independent experiments are shown as means ± SEM. \**P* < 0.05, one-sample t-test. (D) Raltegravir (RAL) or (E) Cabotegravir (CAB) susceptibility of the NC mutants identified in the A5381-Hakim study, measured as described in (B). (F-G) Correlation between fold-change in DTG IC_50_s conferred by NC mutations and fold-change in (F) RAL or (G) CAB IC_50_s. Fold changes in mean INSTI IC_50_ values for each NC mutant, determined by the TZM-bl assay, are plotted. Data include 24 NC mutants (23 single mutants and NC-A30T/M46I) and WT. Of the 24 changes in susceptibility profiles, 13 were previously reported in Hikichi *et al.* (open circle) and 11 (closed circle, including previously described NC-N27I and M46I) were NC mutations that were selected in A5381-Hakim study participants at the time of confirmed virologic failure (CVF). ^24^ Correlations were assessed using a simple linear regression model.

**Table 2.** Susceptibility and Infectivity of HIV-1 with Mutations in Nucleocapsid and/or Integrase against Dolutegravir (DTG)

| Genotype | DTG nM IC<sub>50</sub><sup>a</sup> | Fold-Increase<sup>b</sup> | Infectivity<sup>c</sup> |
| --- | --- | --- | --- |
| WT NL4-3 | 0.78 ± 0.067 | 1 | 1.0 |
| NC-V13I | 0.92 ± 0.062 | 1.2 | 0.86 |
| NC-K20R | 1.2 ± 0.10 | 1.5* | 1.1 |
| NC-N27I | 1.8 ± 0.23 | 2.3* | 1.0 |
| NC-A30T | 1.8 ± 0.057 | 2.3* | 0.99 |
| NC-K34R | 0.99 ± 0.076 | 1.2 | 0.98 |
| NC-K38R | 0.84 ± 0.069 | 1.1 | 1.0 |
| NC-K41N | 1.7 ± 0.12 | 2.2* | 0.68 |
| NC-K41R | 1.1 ± 0.21 | 1.4 | 1.0 |
| NC-Q45R | 1.1 ± 0.21 | 1.5 | 0.90 |
| NC-M46I | 1.3 ± 0.13 | 1.7* | 0.98 |
| NC-A30T/M46I | 2.3 ± 0.13 | 2.9* | 1.2 |
| IN-M50I | 0.92 ± 0.17 | 1.2 | 0.87 |
| IN-H51Y | 0.86 ± 0.21 | 1.1 | 0.91 |
| IN-G118R | 11 ± 2.0 | 14* | 0.40 |
| IN-E157Q | 0.94 ± 0.22 | 1.2 | 0.70 |
| IN-R263K | 2.5 ± 0.39 | 3.2* | 0.65 |
| IN-M50I/R263K | 2.4 ± 0.6 | 3.1* | 0.67 |
| NC-N27I/IN-M50I | 2.0 ± 0.21 | 2.5* | 0.93 |
| NC-N27I/IN-R263K | **5.3 ± 0.4** | 6.8* | 0.75 |
| NC-N27I/IN-M50I/R263K | **6.0 ± 0.71** | 7.6* | 0.87 |
| NC-A30T/M46I/IN-H51Y | 2.6 ± 0.43 | 3.3* | 0.88 |
| RT-M184V | 1.1 ± 0.23 | 1.4 | 1.2 |
| NC-N27I/RT-M184V | 2.7 ± 0.62 | 3.4* | 1.2 |

<sup>a</sup>. IC<sub>50</sub>, mean ± standard error of the mean (SEM) was determined for wild-type (WT) and mutant HIV-1<sub>NL4-3</sub> in TZM-bl cells as described in the Methods. **Bolded IC<sub>50</sub>s** indicate additive or greater fold-increase compared with NC or IN mutations alone.
<sup>b</sup>. Fold-increase was calculated by dividing the IC<sub>50</sub> generated for each mutant by the IC<sub>50</sub> of WT HIV-1<sub>NL4-3</sub>. These data are derived from a minimum of three independent experiments. \**P* < 0.05, determined using one-way analysis of variance (ANOVA), followed by two-stage step-up method of Benjamini, Krieger, and Yekutieli.<sup>57</sup>
<sup>c</sup>. Cell-free infectivity was calculated as relative light units in TZM-bl cells for each mutant divided by the relative light units in TZM-bl cells for WT HIV-1<sub>NL4-3</sub>.

### Phenotypic effects of IN mutations in combination with NC mutations on INSTI resistance

IN mutations alone and in combination with NC mutations observed *in vivo* were evaluated for their phenotypic effect on DTG. The IN-G118R mutation alone conferred the highest level of resistance to DTG (14-fold, p < 0.05). The IN-R263K mutation alone conferred a moderate level of resistance to DTG (3.2-fold, p < 0.05) compared to WT HIV-1_NL4-3_, which increased to 6.8-fold when combined with NC-N27I (p < 0.05) (PID 21). Similarly, the combination of NC-A30T/M46I with IN-H51Y (PID 27) showed a greater reduction in susceptibility to DTG (3.3-fold; p < 0.05) compared to any of the mutations alone (NC-A30T 2.3-fold, p <0.05; NC-M46I 1.7-fold, p <0.05; IN-H51Y no significant effect) **(Table 2**, **Figure 3B)**. Only the IN-G118R and IN-E157Q mutations caused a significant reduction in cell-free infectivity relative to WT HIV-1_NL4-3,_ indicating that reduced susceptibility to DTG from NC and NC with IN mutations was not due to lower cell-free infectivity **(Figure 3C)**.

### Effect of IN/NC mutations on multi-round HIV-1 replication

The mutation combination NC-N27I/IN-R263K (PID 21) was explored further for the effects on multi-round HIV-1 replication. Because IN-M50I was also selected in PID 21, it was included in the genotype (**Supplementary Table S3)**. In the absence of drug, all mutant viruses (NC mutations alone, IN mutations alone, or NC + IN) had equivalent reverse transcriptase (RT) activity in the SupT1 T-cell line over 18 days of infection. Viruses containing both NC-N27I and IN-M50I/R263K mutations were able to replicate in the presence of up to 10 nM DTG and conferred 7.6-fold resistance relative to WT **(Figures 4A and B)**. Consistent with the findings in the single-round TZM-bl assay, evaluation of the mutations observed in PID 21 in primary CD4+ T cells using a single-round VSV-G-pseudotyped nanoLuc reporter assay showed that the combined NC and IN mutations N27I and R263K additively and significantly increased phenotypic resistance, resulting in a 4.8-fold increase in IC_50_ compared with WT **(Figure 4C)**.

**Figure 4.**
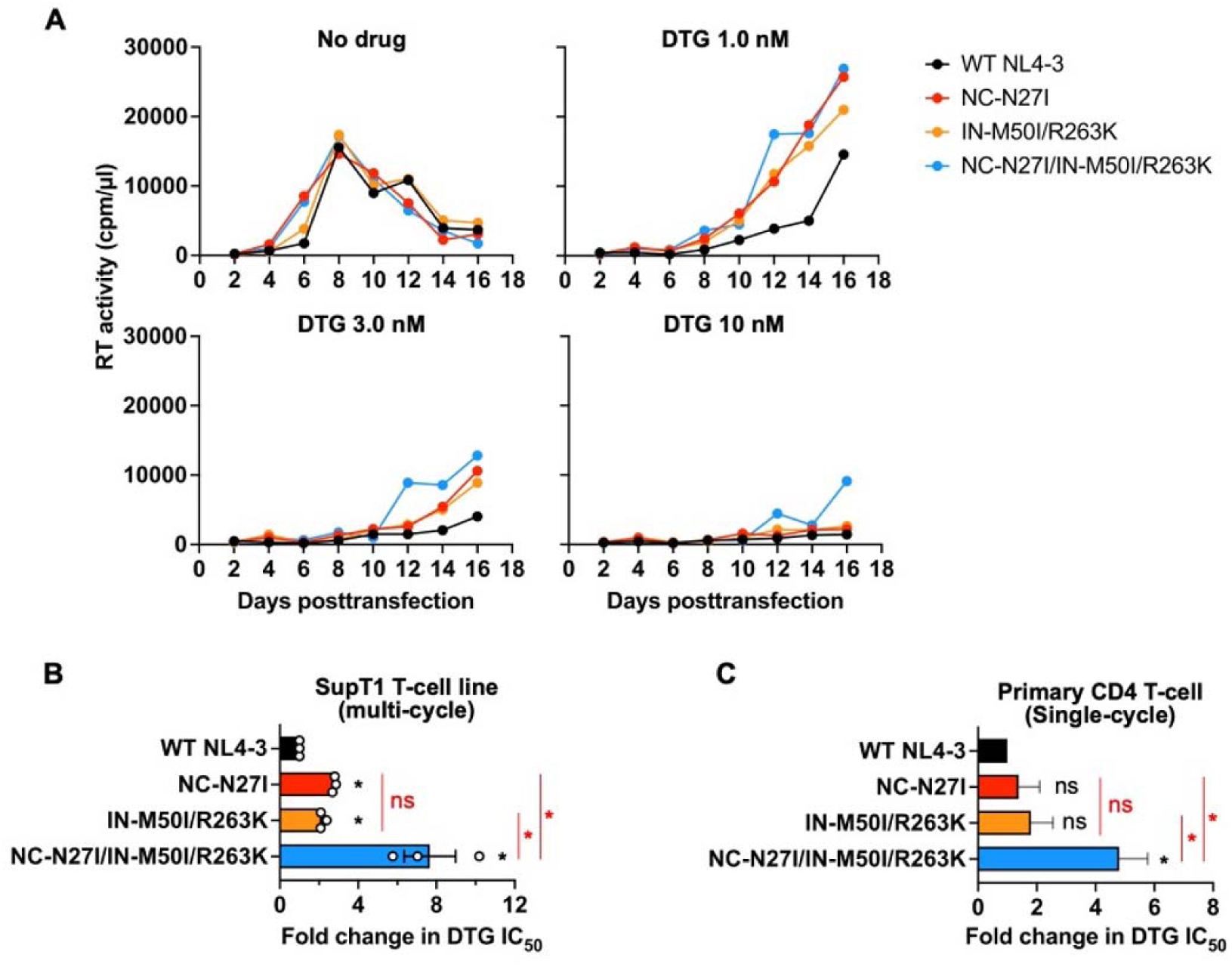
Effect of the NC/IN Mutations Identified in Participant 21 on INSTI susceptibility. (A) Replication kinetics of the indicated NC/IN mutants in SupT1 cells in the presence of DTG (0–1000 nM, 3-fold serial dilutions). Viral replication was monitored longitudinally (RT activity), and representative replication curves at 0, 1, 3, and 10 nM DTG are shown. (B) Fold changes in IC_50_ relative to WT, calculated from the replication kinetics assay shown in (A). For each DTG concentration (0–1000 nM, 3-fold serial dilutions), the area under the replication curve (AUC) was calculated. The IC_50_ was defined as the DTG concentration that reduced the AUC by 50% relative to the no-drug control. Fold change values were determined relative to WT IC_50_. Data shown are means ± SEM from at least three independent experiments. *P < 0.05, one-sample t-test or one-way ANOVA followed by the two-stage step-up method of Benjamini, Krieger, and Yekutieli. ^56^ (C) Susceptibility of NC/IN mutants to DTG in primary CD4+ T cells. Primary CD4+ T cells from four healthy donors were infected with RT-normalized VSV-G–pseudotyped nanoLuc reporter HIV-1 in the presence of increasing concentrations of DTG. Nano-luciferase activity was measured 48 h post-infection. Data are shown as means ± SEM. Statistical significance (*P < 0.05) was determined by one-sample t-test or one-way ANOVA followed by the two-stage step-up method of Benjamini, Krieger, and Yekutieli. ^56^

## DISCUSSION

The A5381-Hakim observational cohort study showed that in ART-naïve individuals starting TLD, or in virologically non-suppressed individuals switching to TLD from NNRTI or PI-based ART, VF with known DTG-associated mutations occurred infrequently, and concentrations of tenofovir-diphosphate (TFV-DP) were lower compared to individuals that did not experience failure, indicating that VF was often associated with suboptimal TLD adherence. ^23,36^ However, some individuals experiencing VF on TLD had no or few major INSTI resistance mutations despite evidence of adherence to TLD based on therapeutic TFV-DP levels in dried blood spots. We therefore investigated if non-canonical mechanisms of INSTI resistance that have been observed *in vitro* could have contributed to VF on TLD by performing whole genome sequencing of plasma HIV-1 RNA before the switch to TLD and at VF on TLD. We found that 11 of 56 individuals with confirmed VF had emergent NC mutations that reduced susceptibility to DTG as determined by *in vitro* IC_50_ determinations of clones encoding the NC mutations. This provides the first clinical evidence that NC mutations emerge with VF of INSTI-containing ART and that the mutations selected decrease susceptibility to INSTI in laboratory assays. Selection of mutations at VF that reduce inhibitor activity are likely to be contributing to the breakthrough viral replication.

Forced-evolution studies from the Freed lab have shown that mutations in off-target genes can reduce susceptibility to DTG in cell culture, leading to the identification of mutations in the NC domain of *gag* that confer decreased susceptibility to DTG. In cell culture, mutations in NC accelerate the kinetics of viral DNA integration, limiting the time for INSTIs to bind to intasomes and block integration ^24,29–31^. Of particular interest are mutations in the NC zinc-finger domain, which span amino acids 13 to 51 and comprise two highly conserved zinc finger motifs that are important for genome recognition, packaging and strand transfer reactions. Mutations in the zinc finger motif of NC could also alter INSTI binding or efficacy during viral DNA integration, without changes in the IN protein itself. ^52–54^

In the current study, 11 of 56 individuals with confirmed VF at or after 6 months of TLD treatment selected one or more mutations (15 alleles total) in the zinc-finger domain of NC that were either undetectable (<5%) pre-TLD (10 alleles) or increased from <20% mutation frequency pre-TLD (5 alleles, mean 13% allele frequency) to ≥20% (mean 57% allele frequency) post-TLD. Notably, zinc finger domain mutations that conferred significantly (p < 0.05) decreased susceptibility to DTG *in vitro* in TZM-bl phenotyping studies were observed in individuals with TFV-DP concentrations above the limit of quantification (NC mutations K41N, N27I, and A30T/M46I in PIDs 20, 21, and 27, respectively), whereas mutations that did not confer decreased susceptibility to DTG *in vitro* were observed in individuals that had TFV-DP levels below the limit of quantification (NC mutation K41R in PIDS 1 and 18, and K34R in PID 56). In three of the 11 individuals, NC mutations emerged in combination with mutations in IN. In all three, TFV-DP levels at VF were in the therapeutic range. *In vitro* analyses of the NC and IN mutations alone and in combination showed that the combined mutations reduced susceptibility to INSTI to a greater extent than individual mutations. The emergence of NC and IN mutations at VF that further reduce INSTI susceptibility provides additional evidence that non-canonical NC mutations can contribute to breakthrough viral replication and treatment failure. Further characterization of which specific NC zinc finger motif mutations or mutation combinations are selected with drug exposure and contribute to reduced INSTI susceptibility will help define signature mutations that could support expanding genotype analysis to include *gag* for clinical management.

One potential concern is whether the sequencing methods used under-detected mutations in IN. We verified sensitive detection of IN mutations using a laboratory-developed next generation sequencing-based whole HIV-1 RNA genome sequencing using mutant:wildtype viral mixture panels to ensure that low-frequency IN mutations at or above 5% frequency were detected. Major INSTI-resistance-conferring mutations IN-G118R and IN-R263K were detected as majority viral variants by both Sanger and WGS. WGS detected few instances of accessory mutations present at frequencies below 20%, including IN-H51Y (5 and 12%) and IN-T97A (9%). These accessory mutations alone are unlikely to explain VF, as IN-T97A was present at study entry and not confirmed VF in one participant (PID 24), and the selection of IN-H51Y at failure in two other participants was at very modest levels (5 to 12% mutation frequency).

As reported previously, IN-R263K conferred 3.2-fold resistance to DTG, which did not change with the addition of IN-N50I (3.1-fold), but increased to 6.8-fold with the addition of NC-N27I as observed in PID 21. The combined interaction between NC-N27I and IN-R263K again highlights NC mutations as amplifiers of DTG resistance, with implications for how VF on TLD is evaluated and managed.

TLD adherence estimated by TFV-DP levels in dried blood spots was generally low in this cohort of individuals who failed TLD. In addition, this analysis only included TFV-DP concentrations at the time of the RNA measurement for confirmed VF, which provides only a single timepoint for adherence estimation. Longitudinal TFV-DP measures are likely to estimate ART adherence more accurately. Exploring additional cohorts, including those with better adherence determined by more frequent measurements, may provide additional evidence for the role of non-canonical mutations in VF of ART.

In summary, this work provides the first clinical evidence of selection of NC mutations with VF of TLD, and of the greater effect of combined selection of NC and IN mutations in reducing susceptibility to DTG. Whether Env mutations, that have also been selected *in vitro* by INSTI, are similarly selected *in vivo* in individuals on failing TLD remains to be determined. ^55^ The findings from this study highlight that HIV-1 can exploit non-canonical mutation pathways to breakthrough INSTI-containing ART and that expanding the regions of that HIV-1 genome analyzed can advance the mechanistic understanding of ART failure.

## Data Availability

Data as presented in this manuscript will be available upon request to the Advancing Clinical Therapeutics Globally (ACTG) and with the written agreement of the ACTG.

## Acknowledgements

This work was presented in part at the 2025 Conference on Retroviruses and Opportunistic Infections, San Francisco, CA March 9-12, 2025. The authors thank the study participants and A5381-Hakim Study Team members.

## Data sharing

Data will be available upon request to the Advancing Clinical Therapeutics Globally (ACTG) and with the written agreement of the ACTG.

## Author Contributions

KP, YH, EF, JM, UP contributed to the conception and design of the work, data analysis, and interpretation. KP, YH, RS, BG, LS, SV, CM, EF, JM, UP contributed to data acquisition and interpretation. CW, EW, CK, CF, MH, SK, and JM provided A5381-Hakim study leadership, direction and oversight. All authors critically reviewed the work for intellectual content and gave final approval for submission. All authors had full access to all the data in the study and agree to be accountable for all aspects of the work. KP and UP directly accessed and verified the data.

## REFERENCES

1. Fairhead C, Levi J, Hill A. Challenges for Novel Antiretroviral Development in an Era of Widespread tenofovir-disoproxil/lamivudine (or emtricitabine)/dolutegravir availability (TLD) Availability. Clin Infect Dis 2025; 80(2): 434–43.

2. World Health Organization. Consolidated Guidelines on HIV Prevention, Testing, Treatment, Service Delivery and Monitoring: Recommendations for a Public Health Approach. WHO; 2021. https://www.who.int/publications/i/item/9789240031593 (accessed June 22, 2026).

3. Cruciani M, Parisi SG. Dolutegravir based antiretroviral therapy compared to other combined antiretroviral regimens for the treatment of HIV-infected naive patients: A systematic review and meta-analysis. PLoS One 2019; 14(9): e0222229.

4. Kanters S, Vitoria M, Zoratti M, et al. Comparative efficacy, tolerability and safety of dolutegravir and efavirenz 400mg among antiretroviral therapies for first-line HIV treatment: A systematic literature review and network meta-analysis. EClinicalMedicine 2020; 28: 100573.

5. Snedecor SJ, Radford M, Kratochvil D, Grove R, Punekar YS. Comparative efficacy and safety of dolutegravir relative to common core agents in treatment-naive patients infected with HIV-1: a systematic review and network meta-analysis. BMC Infect Dis 2019; 19(1): 484.

6. Allahna E, Nicole D, Neha S, et al. Brief Report: Virologic Impact of the Dolutegravir Transition: Prospective Results From the Multinational African Cohort Study. J Acquir Immune Defic Syndr 2022; 91(3): 285–9.

7. Majula RT, Mweya CN. Brief communication: Long-term treatment outcomes of transitioning to dolutegravir-based ART from efavirenz in HIV study participants in Mbeya, Tanzania. AIDS Res Ther 2024; 21(1): 98.

8. Schramm B, Temfack E, Descamps D, et al. Viral suppression and HIV-1 drug resistance 1 year after pragmatic transitioning to dolutegravir first-line therapy in Malawi: a prospective cohort study. Lancet HIV 2022; 9(8): e544–e53.

9. Meireles MV, Pascom ARP, Duarte EC, McFarland W. Comparative effectiveness of first-line antiretroviral therapy: results from a large real-world cohort after the implementation of dolutegravir. AIDS 2019; 33(10): 1663–8.

10. World Health Organization. HIV Drug Resistance: brief report 2024. WHO; 2024. https://www.who.int/publications/i/item/9789240086319 (accessed June 22, 2026).

11. Kingwara L, Onwonga VM, Madada RS, et al. DTG Resistance in Patients with Previous ARV Experience and Viremia in Kenya Receiving DTG-Based ART. Abstract 677. Conference on Retroviruses and Opportunistic Infections. Denver, Colorado; 2024.

12. Chu C, Tao K, Kouamou V, et al. Prevalence of Emergent Dolutegravir Resistance Mutations in People Living with HIV: A Rapid Scoping Review. Viruses 2024; 16(3):399.

13. Henegar C, Letang E, Wang R, et al. A Comprehensive Literature Review of Treatment-Emergent Integrase Resistance with Dolutegravir-Based Regimens in Real-World Settings. Viruses 2023; 15(12):2426.

14. Tao K, Rhee SY, Chu C, et al. Treatment Emergent Dolutegravir Resistance Mutations in Individuals Naive to HIV-1 Integrase Inhibitors: A Rapid Scoping Review. Viruses 2023; 15(9):1932.

15. Castagna A, Maggiolo F, Penco G, et al. Dolutegravir in antiretroviral-experienced patients with raltegravir- and/or elvitegravir-resistant HIV-1: 24-week results of the phase III VIKING-3 study. J Infect Dis 2014; 210(3): 354–62.

16. Loosli T, Hossmann S, Ingle SM, et al. HIV-1 drug resistance in people on dolutegravir-based antiretroviral therapy: a collaborative cohort analysis. Lancet HIV 2023; 10(11): e733–e41.

17. van Osch JAT, Mesplede T. How frequent is dolutegravir resistance? Expert Rev Anti Infect Ther 2025; 23(9): 671–81.

18. Llibre JM, Jou A, Puig T. High-level Dolutegravir Resistance Selection on Dolutegravir/Lamivudine. Clin Infect Dis 2024; 79(6): 1540–1.

19. Rhee SY, Grant PM, Tzou PL, et al. A systematic review of the genetic mechanisms of dolutegravir resistance. J Antimicrob Chemother 2019; 74(11): 3135–49.

20. Tsiang M, Jones GS, Goldsmith J, et al. Antiviral Activity of Bictegravir (GS-9883), a Novel Potent HIV-1 Integrase Strand Transfer Inhibitor with an Improved Resistance Profile. Antimicrob Agents Chemother 2016; 60(12): 7086–97.

21. Quashie PK, Mesplede T, Han YS, et al. Characterization of the R263K mutation in HIV-1 integrase that confers low-level resistance to the second-generation integrase strand transfer inhibitor dolutegravir. J Virol 2012; 86(5): 2696–705.

22. Charpentier C, Descamps D. Resistance to HIV Integrase Inhibitors: About R263K and E157Q Mutations. Viruses 2018; 10(1):41.

23. Marc JB, McCarthy C, Wallis CL, et al. Virological and drug-resistance outcomes for people living with HIV initiating or switching to tenofovir, lamivudine, and dolutegravir in six PEPFAR-supported countries: a prospective cohort study. Lancet HIV 2025; 12(12): e836–e49.

24. Hikichi Y, Grover JR, Schafer A, Mothes W, Freed EO. Epistatic pathways can drive HIV-1 escape from integrase strand transfer inhibitors. Sci Adv 2024; 10(9): eadn0042.

25. Dekker JG, Klaver B, Berkhout B, Das AT. HIV-1 3’-Polypurine Tract Mutations Confer Dolutegravir Resistance by Switching to an Integration-Independent Replication Mechanism via 1-LTR Circles. J Virol 2023; 97(5): e0036123.

26. Malet I, Delelis O, Nguyen T, et al. Variability of the HIV-1 3’ polypurine tract (3’PPT) region and implication in integrase inhibitor resistance. J Antimicrob Chemother 2019; 74(12): 3440–4.

27. Richetta C, Subra F, Malet I, et al. Mutations in the 3’-PPT Lead to HIV-1 Replication without Integration. J Virol 2022; 96(14): e0067622.

28. Richetta C, Tu NQ, Delelis O. Different Pathways Conferring Integrase Strand-Transfer Inhibitors Resistance. Viruses 2022; 14(12):2591.

29. Hikichi Y, Burdick RC, Patro SC, et al. Elucidating the mechanism by which HIV-1 nucleocapsid mutations confer resistance to integrase strand transfer inhibitors. Sci Adv 2025; 11(37): eadz8980.

30. Van Duyne R, Kuo LS, Pham P, Fujii K, Freed EO. Mutations in the HIV-1 envelope glycoprotein can broadly rescue blocks at multiple steps in the virus replication cycle. Proceedings of the National Academy of Sciences of the United States of America 2019; 116(18): 9040–9.

31. Hikichi Y, Van Duyne R, Pham P, et al. Mechanistic Analysis of the Broad Antiretroviral Resistance Conferred by HIV-1 Envelope Glycoprotein Mutations. mBio 2021; 12(1):e03134–20.

32. Le Hingrat Q, Montrouge T, Bachelard A, et al. Nucleocapsid Mutations During Virologic Failure of an Integrase Inhibitor-Based Regimen. Abstract 585. Conference of Retroviruses and Opportunistic Infections. Denver, Colorado; 2026.

33. Coetzee N, Delaney KE, Jennings L, et al. Noncanonical Resistance Mutations in TLD-Treated Individuals with Unexpected Virologic Failure. Abstract 553. Conference on Retroviruses and Opportunistic Infections. Denver, Colorado; 2026.

34. Ngueko AMK, Marchegiani G, Carioti L, et al. Characterization of Mutations Outside the Integrase in HIV-1 Individuals in Whom Dolutegravir Failed. Abstract 550. Conference on Retroviruses and Opportunistic Infections. Denver, Colorado; 2026.

35. Penrose KJ, Hikichi Y, Sethi R, et al. Selection of nucleocapsid mutations with virologic failure of tenofovir-lamivudine-dolutegravir. Abstract 156. Conference on Retroviruses and Opportunistic Infections. San Francisco, CA; 2025.

36. Kityo C, McCarthy C, Koenig SP, et al. Virology Outcomes of Tenofovir-lamivudine-dolutegravir in Treatment-naive and Virologically Suppressed Individuals Switching From an NNRTI-based Regimen: An Observational Analysis at 13 Sites. Open Forum Infect Dis 2025; 12(7): ofaf270.

37. Wallis CL, McCarthy C, Godfrey C, et al. ACTG 5381: Virologic and Resistance Outcomes after Switch to TLD for Failing 1st and 2nd Line ART. Abstract 675. Conference on Retroviruses and Opportunistic Infections (CROI) 2024. Denver, Colorado; 2024.

38. van Heerden JK, Meintjes G, Barr D, et al. Relationship Between Tenofovir Diphosphate Concentrations in Dried Blood Spots and Virological Outcomes After Initiating Tenofovir-Lamivudine-Dolutegravir as First-Line or Second-Line Antiretroviral Therapy. J Acquir Immune Defic Syndr 2024; 95(3): 260–7.

39. Anderson PL, Liu AY, Castillo-Mancilla JR, et al. Intracellular Tenofovir-Diphosphate and Emtricitabine-Triphosphate in Dried Blood Spots following Directly Observed Therapy. Antimicrob Agents Chemother 2018; 62(1): e01710–17.

40. Castillo-Mancilla JR, Zheng JH, Rower JE, et al. Tenofovir, emtricitabine, and tenofovir diphosphate in dried blood spots for determining recent and cumulative drug exposure. AIDS Res Hum Retroviruses 2013; 29(2): 384–90.

41. Yager J, Castillo-Mancilla J, Ibrahim ME, et al. Intracellular Tenofovir-Diphosphate and Emtricitabine-Triphosphate in Dried Blood Spots Following Tenofovir Alafenamide: The TAF-DBS Study. J Acquir Immune Defic Syndr 2020; 84(3): 323–30.

42. Liu TF, Shafer RW. Web resources for HIV type 1 genotypic-resistance test interpretation. Clin Infect Dis 2006; 42(11): 1608–18.

43. Wallis CL, Papathanasopoulos MA, Lakhi S, et al. Affordable in-house antiretroviral drug resistance assay with good performance in non-subtype B HIV-1. Journal of Virological Methods 2010; 163(2): 505–8.

44. Penrose KJ, Wallis CL, Brumme CJ, et al. Frequent Cross-Resistance to Dapivirine in HIV-1 Subtype C-Infected Individuals after First-Line Antiretroviral Therapy Failure in South Africa. Antimicrob Agents Chemother 2017; 61(2): e01805–16.

45. Gall A, Ferns B, Morris C, et al. Universal amplification, next-generation sequencing, and assembly of HIV-1 genomes. J Clin Microbiol 2012; 50(12): 3838–44.

46. Korbie DJ, Mattick JS. Touchdown PCR for increased specificity and sensitivity in PCR amplification. Nat Protoc 2008; 3(9): 1452–6.

47. Mohamed S, Boulme R, Sayada C. From Capillary Electrophoresis to Deep Sequencing: An Improved HIV-1 Drug Resistance Assessment Solution Using In Vitro Diagnostic (IVD) Assays and Software. Viruses 2023; 15(2): 571.

48. Adachi A, Gendelman HE, Koenig S, et al. Production of acquired immunodeficiency syndrome-associated retrovirus in human and nonhuman cells transfected with an infectious molecular clone. J Virol 1986; 59(2): 284–91.

49. Yee JK, Friedmann T, Burns JC. Generation of high-titer pseudotyped retroviral vectors with very broad host range. Methods Cell Biol 1994; 43 Pt A: 99–112.

50. Platt EJ, Wehrly K, Kuhmann SE, Chesebro B, Kabat D. Effects of CCR5 and CD4 cell surface concentrations on infections by macrophagetropic isolates of human immunodeficiency virus type 1. J Virol 1998; 72(4): 2855–64.

51. Malet I, Subra F, Charpentier C, et al. Mutations Located outside the Integrase Gene Can Confer Resistance to HIV-1 Integrase Strand Transfer Inhibitors. mBio 2017; 8(5): 2855–64.

52. Guo J, Wu T, Anderson J, et al. Zinc finger structures in the human immunodeficiency virus type 1 nucleocapsid protein facilitate efficient minus- and plus-strand transfer. J Virol 2000; 74(19): 8980–8.

53. Levin JG, Mitra M, Mascarenhas A, Musier-Forsyth K. Role of HIV-1 nucleocapsid protein in HIV-1 reverse transcription. RNA Biol 2010; 7(6): 754–74.

54. Summers MF, Henderson LE, Chance MR, et al. Nucleocapsid zinc fingers detected in retroviruses: EXAFS studies of intact viruses and the solution-state structure of the nucleocapsid protein from HIV-1. Protein Sci 1992; 1(5): 563–74.

55. Hikichi Y, Penrose KJ, Mathur S, Ablam SD, Parikh UM, Freed EO. Env Mutations Are a Gateway for Acquisition of InSTI Resistance. Abstract 551. Conference on Retroviruses and Opportunistic Infections. Denver, CO; 2026.

56. Benjamini Y, Krieger AM, Yekutieli D. Adaptive linear step-up procedures that control the false discovery rate. Biometrika 2006; 93(3): 491–507.

