## Supplemental Table for "Selection of Mutations in HIV-1 Nucleocapsid and Integrase in Individuals Living with HIV Experiencing Virologic Failure After Initiating or Switching to Tenofovir-Lamivudine-Dolutegravir"

### Appendix
### Table of Contents


| | Page |
|---|---|
| Supplementary Table S1. Local Ethics Committees and National Regulatory Agency Approvals for the ACTG A5381-Hakim Study | 2 |
| Supplementary Table S2. Primary RT-PCR One-Step and Optional Nested PCR Primers for Second Round PCR | 4 |
| Supplementary Table S3. Mutations in Protease, Reverse Transcriptase, Integrase and Nucleocapsid Identified by Whole HIV-1 RNA Next Generation Sequencing in Individuals in A5381 on Failing Tenofovir-Dolutegravir-Lamivudine Antiretroviral Therapy | 5 |

**Supplementary Table S1. Local Ethics Committees and National Regulatory Agency Approvals for the ACTG A5381-Hakim Study**

The ACTG A5381-Hakim Study was approved by the following local ethics committees and national regulatory agencies in the respective countries:

| Site | Name of the Ethics Committee/Institutional Review Board | Decision | Approval Number |
|---|---|---|---|
| Haitian Group for the Study of Kaposi's Sarcoma and Opportunistic Infections | Comite des Droits Humains des Centres GHESKIO (CDH - GHESKIO),<br>Weill-Cornell Medical College (WCMC) IRB | Approved | GHESKIO; 00005515, 00000093 |
| Joint Clinical Research Centre, Kampala Clinical Research Site | Joint Clinical Research Centre / Research Ethics Committee (JCRC / REC),<br>Uganda National Council for Science and Technology (UNCST),<br>University Hospital Cleveland Medical Center (UHCMC) Institutional Review Board | Approved | CRS; JC1019 |
| Kenya Medical Research Institute–Walter Reed Project Clinical Research Center Clinical Research Site | Kenya Medical Research Institute Scientific and Ethics Review Committee Unit (KEMRI SERU),<br>Walter Reed Army Institute of Research (WRAIR) | Approved | SERU 3912, WRAIR 2693 |
| University of the Witwatersrand Clinical HIV Research Unit, Wits Helen Joseph Hospital Clinical Research Site | Witswatersrand Health Research Ethics Committee (WITS HREC),<br>National Health Research Ethics Committee (NHREC) | Approved | 190901 |
| Moi University Clinical Research Centre | Institutional Research Ethics Committee (IREC),<br>National Council for Science and Technology (NCST) | Approved | IREC/2019/000082 |
| Durban International Clinical Research Site, Enhancing Care Foundation | KwaZulu-Natal Department of Health (KZN DOH), Pharma Ethics,<br>Hospital Management | Approved | M19/07/017 |
| University of Cape Town Lung Institute Clinical Research Site | University of Cape Town, Faculty of Health Sciences, Human Research, Ethics Committee (UCT-FHS-HREC) | Approved | 532/2019 |
| Milton Park Clinical Research Site | Medical Research Council of Zimbabwe / Research Council of Zimbabwe (MRCZ/RCZ) | Approved | MRCZ/A/2508 |
| Perinatal HIV Research Unit, University of the Witwatersrand–Soweto Clinical Research Site | Witswatersrand Health Research Ethics Committee (WITS HREC) | Approved | 190901 |

| | | | |
|---|---|---|---|
| Blantyre Clinical Research Site | College of Medicine Research Ethics Committee (COMREC),<br>Johns Hopkins Bloomberg School of Public Health (JHSPH) Institutional Review Board | Approved | P.11/19/2873, 00009752/CR349 |
| University of North Carolina at Chapel Hill Project–Malawi Clinical Research Site | National Health Sciences Research Committee (NHSRC), Pharmacy, Medicines and Poisons Board (PMPB), University of North Carolina (UNC) Institutional Review Board | Approved | NHSRC 19/05/2330, UNC 19-1443 |
| Family Centre for Research with Ubuntu Clinical Research Site | Stellenbosch Health Research Ethics Committee (HREC) | Approved | M19/07/017 |

**Supplementary Table S2. Primary RT-PCR One-Step and Optional Nested PCR Primers for Second Round PCR**

| Primer Type | HIV-1$_{HXB2}$ binding location | Sequence |
|---|---|---|
| **Primary RT-PCR One-Step Primers** | | |
| Amplicon 1-Forward | 456→483 | FWD 5'-GTCTCTCTGGTTAGACCAGATCTGAGCC-3' |
| Amplicon 1-Reverse | 2385←2407 | RVS 5'-CCTCCAATTCCYCCTATCATTTT-3' |
| Amplicon 2-Forward | 1485→1505 | FWD 5'- GGGAAGTGAYATAGCWGGAAC-3' |
| Amplicon 2-Reverse | 5195←5217 | RVS 5'-TAGTGGGATGTGTACTTCTGAAC-3' |
| Amplicon 3-Forward | 4783→4805 | FWD 5'-TTAAAAGAAAAGGGGGGATTGGG-3' |
| Amplicon 3-Reverse | 6858←6880 | RVS 5'-GCACAATAATGTATRGGAATTGG-3' |
| Amplicon 4-Forward | 5967→5985 | FWD 5'-CCTATGGCAGGAAGAAGCG-3' |
| Amplicon 4-Reverse | 9604←9639 | RVS 5'-TAGAGCACTCAAGGCAAGCTTTATTGAGGCTTA-3' |
| **Optional Nested PCR Primers** | | |
| Nested 1-Forward | 513→539 | FWD 5'- ACTGCTTAAGCCTCAATAAAGCTTGCC-3' |
| Nested 1 Reverse | 2366←2392 | RVS-5' ATCATTTTTGGTTTCCATYTTCCTGGC-3' |
| Nested 2 Forward | 2044→2066 | FWD 5'- GAAGGACACCAAATGAAAGAYTG-3' |
| Nested 2 Reverse | 5054←5075 | RVS 5'- TGCCACACAATCATCACCTGCC-3' |
| Nested 3 Foward | 4799→4819 | FWD 5'- GATTGGGGGGTACAGTGCAGG'3' |
| Nested 3 Reverse | 6829←6851 | RVS 5'-GGATACCTTTGGACARGCYTGTG-3' |
| Nested 4 Forward | 5977→5933 | FWD 5'- GAAGAAGCGGAGACARC-3' |
| Nested 4 Reverse | 9497←9517 | RVS 5'- CTTWTATGCAGCWTCTGAGGG-3' |

**Supplementary Table S3. Mutations in Protease, Reverse Transcriptase, Integrase and Nucleocapsid Identified by Whole HIV-1 RNA Next Generation Sequencing in Individuals in A5381 on Failing Tenofovir-Dolutegravir-Lamivudine Antiretroviral Therapy**

**A5381 Study Groups.** Participants in A5381 were enrolled in study groups as follows and as previously described [1,2]: Group 1a included individuals with HIV-1 RNA >1000 copies/mL at study entry who switched from non-nucleoside reverse transcriptase inhibitor (NNRTI)-based first-line antiretroviral therapy (ART) to tenofovir-lamivudine-dolutegravir (TLD). Group 2a included individuals with HIV-1 RNA >1000 copies/mL at study entry who switched from protease inhibitor (PI)-based second-line ART to TLD). Group 3 included individuals receiving first-line TLD with concomitant rifampicin treatment plus an additional 50 mg dolutegravir (DTG). Group 4 included individuals initiating first-line TLD.

**A5381 Analysis-Specific Study Visits.** For this analysis, samples were tested that were collected at A5381 study entry (ENT) prior to TLD initiation. Samples were also collected at the time of confirmed virologic failure (CVF), which was the second of two consecutive follow-up visits with plasma HIV-1 RNA >1000 copies/mL, which could have occurred up to six months after the first visit with HIV-1 RNA >1000 copies/mL.

**Determination of Drug Levels at CVF.** A dried blood spot card (DBS) was collected at the visit at which virologic failure was confirmed to assess for drug levels as a marker for ART adherence. Tenofovir diphosphate (TFV-DP) concentrations were measured at the University of Cape Town from dried blood spots collected at CVF to approximate ART adherence, as previously described. [3] Briefly, a 25 µl DBS punch was extracted with methanol: water and isotopic tenofovir as an internal standard, then TFV-DP was quantified by validated liquid chromatography/tandem mass spectrometry. The lower limit of quantification was 16.6 femtomole (fmol)/3mm punch. [4-6]

**Whole HIV-1 RNA Genome Sequencing (WGS)**. A laboratory-developed whole HIV-1 RNA genome sequencing (HIV-1 WGS) method was used to identify mutations in nucleocapsid (NC), protease (PR), reverse transcriptase (RT) and integrase (IN) at a variant frequency of ≥5%. Total RNA was extracted from plasma using guanidinium thiocyanate lysis followed by isopropanol precipitation. HIV-1 whole-genome Illumina libraries were prepared using an approach adapted from Gall *et al.* to amplify HIV-1 RNA genomes in four overlapping one-step RT-PCR reactions (Superscript IV One Step, ThermoFisher Scientific). [7] Primers were optimized for subtype diversity, optimal amplification across a range of HIV-1 RNA >1000 copies/mL and long terminal repeat (LTR) coverage through partial R and full U5 in amplicon one and full U3 and partial R in amplicon four. Optional nested PCR primers were used in a second round of PCR (SuperFI, ThermoFisher Scientific) for samples with poor amplification **(Supplementary Table S1)**. [8] Amplicons were purified using bead- or agarose gel-based purification (KAPA Pure Bead, Roche) and quantified by PicoGreen Quant-it (Thermo Fisher Scientific) prior to library preparation (PurePlex, SeqWell). Libraries were pooled, quantified, and sequenced on the Illumina MiSeq platform. Paired-end reads were demultiplexed using Illumina software, and FASTQ files were generated for downstream bioinformatic analysis.

**Bioinformatics.** DeepChek® software using a mutation threshold of five percent frequency was used for analysis of all HIV-1 gene targets. Drug resistance mutations (DRMs) were identified by the DeepChek® software using Stanford's HIVdb v9.8 algorithm. [9-12] The frequency of each variant present at ≥5% was obtained from the Amino Acid - Quality Information Report. Mutations were considered selected at confirmed VF on TLD if they were detected at ≥20% frequency at confirmed VF and were either absent at study entry or present at <20% frequency at study entry.

| PID | Gp | Visit (Month)[a] | HIV-1 RNA (copies/ mL) | TFV-DP in DBS (fmol/ 3mm punch)[b] | Subtype | Protease Mutations (% Allele Frequency)[c] | Reverse Transcriptase Mutations (% Allele Frequency)[d] | Integrase Mutations (% Allele Frequency)[e] | Nucleocapsid Mutations (% Allele Frequency)[f] |
|---|---|---|---|---|---|---|---|---|---|
| **1** | 1a | ENT (0) | 1,475 | ·· | C | None | D67N (99), K70R (99), K103N (99), V108I (99), M184V (94), T215I (61), T215V (39), K219E (99), N348I (99) | None | T24I (99) |
| **1** | 1a | CVF (27) | 11,583 | <16.6 | C | None | None | None | T24I (99), **K41R (92)** |
| **2** | 1a | ENT (0) | 4,700 | ·· | A1 | None | K103N (33), K103S (38) | None | V13I (96), T24L (100) |

| PID | Gp | Visit (Month)[a] | HIV-1 RNA (copies/ mL) | TFV-DP in DBS (fmol/ 3mm punch)[b] | Subtype | Protease Mutations (% Allele Frequency)[c] | Reverse Transcriptase Mutations (% Allele Frequency)[d] | Integrase Mutations (% Allele Frequency)[e] | Nucleocapsid Mutations (% Allele Frequency)[f] |
|---|---|---|---|---|---|---|---|---|---|
| **2** | 1a | CVF (28) | 6,491 | <16.6 | A1 | None | K103N (27), K103S (20), V106I (22) | None | V13I (99), T24L (99) |
| **3** | 1a | ENT (0) | 12,258 | ·· | A1 | None | T69G (99), K70R (100), L100I (100), K103N (98), V179T (35), M184V (99), K219E (99), Y318F (16) | None | V13I (83), T24I (8), T24L (92), K34R (23) |
| **3** | 1a | CVF (11) | 21,449 | 18.3 | A1 | None | T69G (41), K70R (16), L100I (41), K103N (97), V179T (11), K219E (17), Y318F (7) | None | V13I (97), T24L (99) |
| **4** | 1a | ENT (0) | 91,921 | ·· | D | None | K65R (53), S68G (36), S68N (18), K70E (5), K70Q (43), V90I (15), V106I (99), M184V (100), Y188L (100), H221Y (18), K238N (25) | None | V13I (96), K41N (7), M46L (100) |
| **4** | 1a | CVF (27) | 4,650 | 79.7 | D | None | M41L (22), K70Q (66), V106I (87), M184V (61), Y188L (100), K238N (42) | None | V13I (98), K41N (18), M46L (100) |
| **5** | 2a | ENT (0) | 33,400 | ·· | B | None | V90I (79), K103R (77) | None | T24I (100), R29K (9), K41R (34) |
| **5** | 2a | CVF (9) | 300,738 | 367 | B | None | V90I (66), K103R (67) | None | T24I (100), K41R (57) |
| **6** | 2a | ENT (0) | 16,168 | ·· | B | K43T (99) | M41L (92), M184V (91), T215F (99) | None | T24I (100) |
| **6** | 2a | CVF (11) | 299,602 | 723 | B | K43T (98) | M41L (65), M184V (65), T215F (99) | **G118R (62)** | T24I (99), **K41G (41)** |
| **7** | 2a | ENT (0) | 45,563 | ·· | B | None | K103N (99) | *L74I (99)* | K20R (7), T24I (100), R26K (92), R29K (5), K34Q (93), K41R (61) |
| **7** | 2a | CVF (9) | 4,306 | 287 | B | None | K103N (99) | *L74I (99)* | T24I (100), R26K (100), K34Q (99), K41R (78) |
| **8** | 2a | ENT (0) | 26,515 | ·· | B | None | D67N (5), K70E (7), V108I (99), V179E (98) | None | T24I (100), K33R (11), K41Q (99), T50P (12), E51G (100) |
| **8** | 2a | CVF (17) | 1,205 | 363 | B | None | D67N (35), K70E (37), K70T (12), V108I (100), V179E (78), T215N (24) | None | K14R (6), T24I (100), A30T (6), K41Q (99), E51G (100) |
| **9** | 2a | ENT (0) | 22,500 | ·· | B | None | K103N (36), K103R (37) | None | T24I (96), R26K (99), K41Q (71) |
| **9** | 2a | CVF (6) | 33,994 | 771 | B | None | K103R (36) | None | T24I (99), R26K (99), K41Q (99) |
| **10** | 2a | ENT (0) | 25,149 | ·· | B | None | K103N (56), K103S (43), V108I (10) | None | V13I (92), E21T (99), T24I (100), R26K (100), R29K (98), D48E (7) |
| **10** | 2a | CVF (10) | 5,649 | <16.6 | B | None | K103N (58), K103S (42) | None | V13I (98), E21T (100), T24I (100), R26K (98), R29K (100), M46I (8) |
| **11** | 2a | ENT (0) | 373,699 | ·· | B | None | None | None | V13I (100), T24I (100), R26K (99), D48E (98) |
| **11** | 2a | CVF (10) | 1,082 | 356 | B | None | None | None | V13I (100), T24I (100), R26K (100), K41R (8), D48E (100) |
| **12** | 2a | ENT (0) | 4,084 | ·· | C | None | S68G (100), A98G (100), K101E (97), M184V (99), G190A (100) | None | T24I (99), R26K (95), T50S (98) |
| **12** | 2a | CVF (11) | 30,983 | <16.6 | C | None | S68G (100), T69D (5), A98G (100), K101E (33), M184V (14), G190A (100) | None | T24I (99), R26K (100), T50S (99) |
| **13** | 2a | ENT (0) | 7,005 | ·· | C | None | K103N (99) | None | T24I (100), K41R (6) |

| PID | Gp | Visit (Month)[a] | HIV-1 RNA (copies/ mL) | TFV-DP in DBS (fmol/ 3mm punch)[b] | Subtype | Protease Mutations (% Allele Frequency)[c] | Reverse Transcriptase Mutations (% Allele Frequency)[d] | Integrase Mutations (% Allele Frequency)[e] | Nucleocapsid Mutations (% Allele Frequency)[f] |
|---|---|---|---|---|---|---|---|---|---|
| **13** | 2a | CVF (11) | 62,077 | 135 | C | None | K103N (99) | None | T24I (100), K41R (7) |
| **14** | 2a | ENT (0) | 50,464 | ·· | A1 | None | E44D (97), S68G (100) | None | T24L (100), E42D (99) |
| **14** | 2a | CVF (6) | 13,734 | 78.9 | A1 | None | E44D (98), S68G (99), V106I (16) | None | T24L (100), E42D (99) |
| **15** | 2a | ENT (0) | 139,834 | ·· | A1 | None | G190A (99) | None | V13I (93), T24L (100) |
| **15** | 2a | CVF (7) | 61,795 | <16.6 | A1 | None | G190A (100) | None | V13I (97), T24L (100) |
| **16** | 2a | ENT (0) | 64,772 | ·· | A1 | K20T (99), M46I (99), I84V (99), L89T (99) | M41L (99), V75M (99), F77L (96), A98G (99), K101E (98), M184V (99), G190A (99), T215Y (98) | None | V13I (97), T24L (99), K41Q (44) |
| **16** | 2a | CVF (7) | 2,004 | 329 | A1 | K20T (99), M46I (100), I84V (100), L89T (100) | M41L (100), V75M (100), F77L (100), A98G (95), K101E (100), M184V (100), G190A (100), T215Y (100) | *H51Y (5)*, **G118R (100)** | V13I (99), T24L (100) |
| **17** | 2a | ENT (0) | 258,254 | ·· | A1 | None | V106I (99), Y188L (99) | *L74I (99)* | V13I (90), T24L (100), R26K (6), K41R (91) |
| **17** | 2a | CVF (9) | 235,834 | 54.2 | A1 | None | V106I (100), Y188L (100) | *L74I (98)* | V13I (97), T24L (100), K41R (97) |
| **18** | 2a | ENT (0) | 3,947 | ·· | A1 | None | M184V (91), Y188C (30), H221Y (30) | None | T24L (99), R26K (99), K41R (7) |
| **18** | 2a | CVF (11) | 44,746 | <16.6 | A1 | None | None | None | T24L (100), R26K (100), **K41R (53)** |
| **19** | 2a | ENT (0) | 179,114 | ·· | A1 | None | None | None | V13I (97), T24L (99), D48E (96) |
| **19** | 2a | CVF (7) | 283,564 | 392 | A1 | None | None | None | V13I (99), T24L (100), D48E (100) |
| **20** | 2a | ENT (0) | 135,666 | ·· | A1 | None | None | None | V13I (100), T24L (99) |
| **20** | 2a | CVF (16) | 31,012 | 211 | D | None | None | None | V13I (99), T24L (98), K34R (13), **K41N (82)** |
| **21** | 2a | ENT (0) | 717,292 | ·· | A1 | None | K65R (100), D67G (37), S68G (100), L100I (99), K103N (100), E138A (100), M184V (100), K219E (100), M230L (100) | None | V13I (75), T24L (100), C49insX (92), T50N (85) |
| **21** | 2a | CVF (8) | 29,854 | 693 | A1 | None | K65R (99), S68G (100), L100I (99), K103N (92), E138A (100), M184V (99), K219E (100), M230L (100) | **R263K (100)** | V13I (90), T24L (98), **N27I (77)**, M46I (16), C49insX (92), T50N (94) |
| **22** | 2a | ENT (0) | 1,153 | ·· | A1 | None | D67G (96), K70E (94), K101E (98), V179F (11), Y181C (62), M184V (100), G190S (100) | None | V13I (99), T24L (99), K41N (99), E51M (46) |
| **22** | 2a | CVF (9) | 5,010 | 360 | A1 | None | D67G (94), K70E (93), K101E (87), V106M (8), Y181C (25), M184V (99), G190S (99) | None | V13I (97), T24L (98), R26K (15), K41N (98), E51M (34) |
| **23** | 2a | ENT (0) | 252,369 | ·· | A1 | None | K103N (17) | *L74I (100)* | T24I (20), T24V (79), N27H (10) |
| **23** | 2a | CVF (7) | 7,179 | 401 | A1 | None | | *L74I (100)* | T24V (95) |
| **24** | 2a | ENT (0) | 1,742 | ·· | A1 | None | E44D (79), S68G (93), A98G (100), Y181C (93), M184V (99), T215Y (100) | *T97A (9)* | T24L (99) |
| **24** | 2a | CVF (17) | 6,924 | 107 | A1 | None | E44D (95), S68G (99), A98G (100), Y181C (99), T215C (21), T215D (20), | None | T24L (99) |

| PID | Gp | Visit (Month)[a] | HIV-1 RNA (copies/ mL) | TFV-DP in DBS (fmol/ 3mm punch)[b] | Subtype | Protease Mutations (% Allele Frequency)[c] | Reverse Transcriptase Mutations (% Allele Frequency)[d] | Integrase Mutations (% Allele Frequency)[e] | Nucleocapsid Mutations (% Allele Frequency)[f] |
|---|---|---|---|---|---|---|---|---|---|
| | | | | | | | T215N (9), T215S (19), T215Y (30) | | |
| **25** | 2a | ENT (0) | 2,680 | ·· | A1 | None | M184V (94), P225H (44) | None | V13I (9), T24L (100), T50N (6), T50S (62) |
| **25** | 2a | CVF (10) | 6,186 | 467 | A1 | None | P225H (15) | None | T24L (100), T50S (97) |
| **26** | 2a | ENT (0) | 4,652 | ·· | A1 | None | None | None | V13I (99), T24L (100) |
| **26** | 2a | CVF (10) | 7,982 | 58.9 | A1 | None | None | None | V13I (100), T24L (100) |
| **27** | 2a | ENT (0) | 56,630 | ·· | A1 | None | E44D (21), S68G (96), K103N (77), K103S (22), V179T (61), T215F (7), T215L (12), K219R (11) | None | T24L (100) |
| **27** | 2a | CVF (28) | 1,929 | 1251 | A1 | None | S68G (99), K70E (8), V90I (17), K103N (99), V179T (6), M184I (6), M184V (86), T215I (6), T215S (11) | *H51Y (12)* | T24L (97), **A30T (59), M46I (69)** |
| **28** | 2a | ENT (0) | 1,685 | ·· | A1 | None | V179T (100) | None | V13I (99), T24L (98) |
| **28** | 2a | CVF (11) | 6,552 | <16.6 | A1 | None | V179T (100) | None | V13I (99), T24L (99) |
| **29** | 2a | ENT (0) | 515,752 | ·· | A1 | None | V179T (100), Y181C (20), H221Y (24) | None | V13I (98), T24I (72), T24L (28), T50insM (93) |
| **29** | 2a | CVF (11) | 150,629 | <16.6 | A1 | None | V179T (100), Y181C (28), H221Y (34) | None | V13I (99), T24I (62), T24L (36), T50insM (95) |
| **30** | 2a | ENT (0) | 5,222 | ·· | A1 | F53L (97) | K103N (99), M184V (99), K238T (18), N348I (100) | None | V13I (100), T24V (97), T50N (99) |
| **30** | 2a | CVF (27) | 2,389 | <16.6 | A1 | F53L (11) | K103N (92) | None | V13I (99), T24V (99), T50N (97) |
| **31** | 2a | ENT (0) | 10,160 | ·· | CRF21_A2D | None | None | None | V13I (96), T24I (100), R26K (100), K41R (51), E42D (25) |
| **31** | 2a | CVF (10) | 57,700 | <16.6 | CRF21_A2D | None | None | None | V13I (100), T24I (100), R26K (99), K41R (27), E42D (18) |
| **32** | 2a | ENT (0) | 63,726 | ·· | A1 | None | A98G (100), Y181C (42) | None | V13I (79), T24L (99), R29K (34), K34R (78), K41R (30) |
| **32** | 2a | CVF (11) | 298,371 | 281 | A1 | None | A98G (100), Y181C (100) | None | V13I (87), T24L (100), R29K (21), K34R (74), K41R (70) |
| **33** | 2a | ENT (0) | 9,959 | ·· | A1 | L10F (63), M46I (12), I84V (97), L89T (100) | K70R (99), K103N (100), V179T (8), M184V (100), K219Q (100), M230L (100), L234I (100) | None | T24L (100), P31L (6) |
| **33** | 2a | CVF (16) | 2,237 | ·· | A1 | L23I (73), L89T (100) | K70R (100), K103N (96), M184V (100), K219Q (100), M230L (100), L234I (99) | *L74M (28),* **G118R (100)** | T24L (100), A25V (9) |
| **34** | 2a | ENT (0) | 43,440 | ·· | B | None | K103N (23) | None | T24I (99), R26K (99), T50N (16), T50insX (55) |
| **34** | 2a | CVF (9) | 219,452 | <16.6 | B | None | | None | T24I (99), R26K (100), T50N (11), T50insX (69) |
| **35** | 2a | ENT (0) | 16,347 | ·· | C | None | K103N (98) | None | T24I (97) |
| **35** | 2a | CVF (23) | 78,130 | <16.6 | C | None | K103N (98) | None | T24I (100) |
| **36** | 2a | ENT (0) | 56,299 | ·· | B | None | Y181C (94), H221Y (91) | None | T24I (99), K34R (99), K41R (100) |

| PID | Gp | Visit (Month)[a] | HIV-1 RNA (copies/ mL) | TFV-DP in DBS (fmol/ 3mm punch)[b] | Subtype | Protease Mutations (% Allele Frequency)[c] | Reverse Transcriptase Mutations (% Allele Frequency)[d] | Integrase Mutations (% Allele Frequency)[e] | Nucleocapsid Mutations (% Allele Frequency)[f] |
|---|---|---|---|---|---|---|---|---|---|
| **36** | 2a | CVF (26) | 169,019 | 180 | B | None | Y181C (99), H221Y (99) | None | E21D (6), T24I (100), **N27S (32)**, K34R (99), K41R (99) |
| **37** | 2a | ENT (0) | 1,471 | ·· | B | None | V179D (75), M184V (99) | None | T24I (99), R26K (79), K34R (61) |
| **37** | 2a | CVF (16) | 50,939 | ·· | B | None | V179D (60), M184V (94) | None | T24I (100), R26K (59), K34R (37) |
| **38** | 2a | ENT (0) | 37,029 | ·· | B | None | K103N (60) | *E157Q (99)* | T24I (99),T50D (60) |
| **38** | 2a | CVF (6) | 24,766 | 41.9 | B | None | K103N (49) | *E157Q (99)* | T24I (99),T50D (46) |
| **39** | 2a | ENT (0) | 236,791 | ·· | B | None | K103N (98) | *E157Q (99)* | V13I (99), T24I (100), R26K (100), R29K (99), K41R (66) |
| **39** | 2a | CVF (15) | 10,112 | ·· | B | None | K103N (97) | *E157Q (99)* | V13I (100), T24I (100), R26K (100), R29K (100), K41R (99) |
| **40** | 2a | ENT (0) | 29,939 | ·· | B | None | S68G (41), V179E (37), K238T (52) | *E157Q (99)* | E21I (86), T24I (99), R26K (100), K34R (8), K41Q (98) |
| **40** | 2a | CVF (8) | 28,224 | 336 | B | None | V179E (97), K238T (98) | *E157Q (99)* | E21I (39), **E21V (38)**, T24I (99), R26K (100), K34R (19), K41Q (99) |
| **41** | 2a | ENT (0) | 17,661 | ·· | B | None | K103R (96) | None | T24I (100), R26K (100), K33R (98), K41R (98) |
| **41** | 2a | CVF (15) | 17,079 | ·· | B | None | K103R (98) | None | T24I (99), R26K (100), K33R (100), K41R (100) |
| **42** | 2a | ENT (0) | 31,736 | ·· | B | None | K103N (16) | *L74I (100), E157Q (100)* | T24I (100), K41Q (98), T50N (83) |
| **42** | 2a | CVF (18) | 4,546 | 455 | B | None | None | *L74I (97), E157Q (100)* | T24I (99), K41Q (99), T50N (93) |
| **43** | 3 | ENT (0) | 545,141 | ·· | C | None | None | None | V13I (96), T24I (100), K41E (36), E51G (34) |
| **43** | 3 | CVF (12) | 1,506,62 1 | <16.6 | C | None | None | None | V13I (99), T24I (100), K41E (77), E51G (75) |
| **44** | 3 | ENT (0) | 262,058 | ·· | C | None | None | None | V13I (100), T24I (98), R26K (100) |
| **44** | 3 | CVF (16) | 25,336 | 207 | C | None | None | None | V13I (100), T24I (100), R26K (100) |
| **45** | 3 | ENT (0) | 381,469 | ·· | C | None | A98G (100), V106M (100), Y181C (100), K219Q (70) | None | T24I (99) |
| **45** | 3 | CVF (17) | 30,091 | <16.6 | C | None | A98G (99), V106M (99), Y181C (99), K219Q (32) | None | T24I (99) |
| **46** | 3 | ENT (0) | 670,147 | ·· | C | None | None | None | V13I (10), T24I (100), K38R (17), K41Q (83), M46I (17) |
| **46** | 3 | CVF (11) | 122,803 | <16.6 | C | None | None | None | **V13I (50),** T24I (99), **K38R (99), M46I (89)** |
| **47** | 4 | ENT (0) | 35,599 | · | B | None | K103N (7) | None | K20R (15), T24I (99), A25S (18), K41R (99) |
| **47** | 4 | CVF (6) | 56,887 | 33.9 | B | None | K103N (7) | None | **K20R (28),** T24I (100), A25S (6), K41R (67), **Q45R (29)** |
| **48** | 4 | ENT (0) | 5,642 | ·· | B | None | K103N (37) | None | V13I (100), T24I (100), R26K (100), T50I (99) |
| **48** | 4 | CVF (11) | 2,158 | 23.8 | B | None | None | None | V13I (100), T24I (100), R26K (100), E42K (7), T50I (100) |

| PID | Gp | Visit (Month)[a] | HIV-1 RNA (copies/ mL) | TFV-DP in DBS (fmol/ 3mm punch)[b] | Subtype | Protease Mutations (% Allele Frequency)[c] | Reverse Transcriptase Mutations (% Allele Frequency)[d] | Integrase Mutations (% Allele Frequency)[e] | Nucleocapsid Mutations (% Allele Frequency)[f] |
|---|---|---|---|---|---|---|---|---|---|
| **49** | 4 | ENT (0) | 241,288 | ·· | B | None | V179D (14) | None | V13I (98), T24I (99), R26K (99), N27H (98), K41R (99) |
| **49** | 4 | CVF (16) | 45,546 | <16.6 | B | None | None | None | V13I (100), T24I (100), R26K (100), N27H (99), K41R (100) |
| **50** | 4 | ENT (0) | 311,712 | ·· | A1 | None | None | *L74I (33)* | V13I (99), T24L (99) |
| **50** | 4 | CVF (10) | 802,467 | ·· | A1 | None | None | *L74I (47)* | V13I (100), T24L (100) |
| **51** | 4 | ENT (0) | 56,925 | ·· | D | None | V90I (35), K103N (98) | None | V13I (99), R26K (99), K41R (98) |
| **51** | 4 | CVF (16) | 38,333 | 97.9 | D | None | V90I (25), K103N (99) | None | V13I (100), R26K (99), K41R (100) |
| **52** | 4 | ENT (0) | 48,864 | ·· | C | None | None | None | T24I (100), K34R (99) |
| **52** | 4 | CVF (12) | 100,886 | <16.6 | C | None | None | None | T24I (100), K34R (99) |
| **53** | 4 | ENT (0) | 266,389 | ·· | C | None | D67N (81), K70E (6), K70Q (22), K103N (76), V106A (17), P225H (43), F227L (44), K238T (17) | None | T24I (99), R26K (100) |
| **53** | 4 | CVF (11) | 339,778 | <16.6 | C | None | None | None | T24I (99), R26K (99) |
| **54** | 4 | ENT (0) | 179,437 | ·· | C | None | None | None | E21A (98), T24I (98), R26K (99) |
| **54** | 4 | CVF (7) | 529,877 | <16.6 | C | None | None | None | E21A (98), T24I (98), R26K (99) |
| **55** | 4 | ENT (0) | 1,262,06 3 | ·· | B | None | None | None | R7K (39), I12A (69), I12T (9), I12V (21), T24I (99) |
| **55** | 4 | CVF (9) | 340,440 | 683 | B | None | None | None | R7K (49), N8S (10), I12A (81), I12T (15), T24I (99) |
| **56** | 4 | ENT (0) | 5,103 | ·· | B | None | K103N (99) | None | T24I (99) |
| **56** | 4 | CVF (10) | 19,604 | <16.6 | B | None | K103N (100) | None | T24I (100), **K34R (23)** |

[a.] Listed in parentheses is the number of months after study entry at which the sample was collected.
[b.] The limit of quantitation (LOQ) for tenofovir-diphosphate (TFV-DP) in DBS was 16·6 femtomoles (fmol) per three millimeter (mm) DBS punch. The double dots "··" indicate that the test was not performed.
[c.] Mutations in protease listed on the table were defined by Stanford HIVdb v9.8 as major mutations. "None" indicates no major drug resistance-associated mutations were detected.
[d.] Mutations in reverse transcriptase were defined by Stanford HIVdb v9.8 as nucleoside or nucleotide reverse transcriptase inhibitor (NRTI) or non-nucleoside reverse transcriptase inhibitor (NNRTI)-associated major mutations. "None" indicates no drug resistance-associated mutations were detected.
[e.] Mutations in integrase were defined by Stanford HIVdb v9.8 as **major (bold)**, *accessory(italics)*, or other. "None" indicates no drug resistance-associated mutations were detected.
[f.] Mutations in the nucleocapsid zinc finger motif (amino acids 13 to 51) were identified by the DeepChek® software by comparison to HIV-$1_{HXB2}$ wild-type reference. Mutations that were selected as defined by detection <20% frequency at study entry and ≥ 20% at the time of confirmed virologic failure are indicated in **bold**.

Abbreviations: Antiretroviral Therapy (ART); Confirmed Virologic Failure Study Visit (CVF); Dried Blood Spot (DBS); Limit of Quantitation (LOQ); Participant identifier (PID); Study Entry Visit (ENT); Study Group (Gp)

**References:**

1. Kityo C, McCarthy C, Koenig SP, et al. Virology Outcomes of Tenofovir-lamivudine-dolutegravir in Treatment-naive and Virologically Suppressed Individuals Switching From an NNRTI-based Regimen: An Observational Analysis at 13 Sites. *Open Forum Infect Dis* 2025; **12**(7): ofaf270.

2. Marc JB, McCarthy C, Wallis CL, et al. Virological and drug-resistance outcomes for people living with HIV initiating or switching to tenofovir, lamivudine, and dolutegravir in six PEPFAR-supported countries: a prospective cohort study. *Lancet HIV* 2025; **12**(12): e836-e49.
3. van Heerden JK, Meintjes G, Barr D, et al. Relationship Between Tenofovir Diphosphate Concentrations in Dried Blood Spots and Virological Outcomes After Initiating Tenofovir-Lamivudine-Dolutegravir as First-Line or Second-Line Antiretroviral Therapy. *J Acquir Immune Defic Syndr* 2024; **95**(3): 260-7.
4. Anderson PL, Liu AY, Castillo-Mancilla JR, et al. Intracellular Tenofovir-Diphosphate and Emtricitabine-Triphosphate in Dried Blood Spots following Directly Observed Therapy. *Antimicrob Agents Chemother* 2018; **62**(1): e01710-17.
5. Castillo-Mancilla JR, Zheng JH, Rower JE, et al. Tenofovir, emtricitabine, and tenofovir diphosphate in dried blood spots for determining recent and cumulative drug exposure. *AIDS Res Hum Retroviruses* 2013; **29**(2): 384-90.
6. Yager J, Castillo-Mancilla J, Ibrahim ME, et al. Intracellular Tenofovir-Diphosphate and Emtricitabine-Triphosphate in Dried Blood Spots Following Tenofovir Alafenamide: The TAF-DBS Study. *J Acquir Immune Defic Syndr* 2020; **84**(3): 323-30.
7. Gall A, Ferns B, Morris C, et al. Universal amplification, next-generation sequencing, and assembly of HIV-1 genomes. *J Clin Microbiol* 2012; **50**(12): 3838-44.
8. Korbie DJ, Mattick JS. Touchdown PCR for increased specificity and sensitivity in PCR amplification. *Nat Protoc* 2008; **3**(9): 1452-6.
9. Mohamed S, Boulme R, Sayada C. From Capillary Electrophoresis to Deep Sequencing: An Improved HIV-1 Drug Resistance Assessment Solution Using In Vitro Diagnostic (IVD) Assays and Software. *Viruses* 2023; **15**(2): 571.
10. Rhee S-Y, Gonzales MJ, Kantor R, et al. Human immunodeficiency virus reverse transcriptase and protease sequence database. *Nucleic Acids Research* 2003; **31**(1), 298-303.
11. Shafer RW. Rationale and Uses of a Public HIV Drug-Resistance Database. *Journal of Infectious Diseases* 2006; **194**(Suppl 1):S51-8
12. Liu TF, Shafer RW. Web Resources for HIV type 1 Genotypic-Resistance Test Interpretation. *Clin Infect Dis* 2006. **42**(11):1608-18.
